# Determinants of COVID-19 booster uptake in the Netherlands, autumn 2022: how well were those at risk for severe disease reached?

**DOI:** 10.1101/2023.08.04.23293632

**Authors:** Caren van Roekel, Lisanne Labuschagne, Joyce Pijpers, Annika van Roon, Bente Smagge, José A. Ferreira, Susan Hahné, Hester de Melker

## Abstract

**Background:** A booster with bivalent COVID-19 vaccine was offered in the Netherlands in autumn, 2022. We aimed to investigate vaccine uptake during the autumn 2022 booster round among the population subgroups at risk for severe COVID-19 that were specifically targeted by this campaign: the medical risk group aged 18-59 years and individuals ≥60 years. We calculated booster uptake in both populations and analyzed determinants of booster uptake among those who had received at least one prior COVID-19 vaccination.

**Methods:** Having had an autumn 2022 booster dose was defined as having received a COVID-19 vaccination between 19 September 2022 and 7 March 2023. The study population included individuals who received at least one previous COVID-19 vaccination. National registries of sociodemographic determinants and COVID-19 vaccination were linked by a unique person identifier. Voting proportions for political parties were included at neighborhood level. Determinants of COVID-19 vaccine autumn booster uptake were ranked by importance by random forest analyses.

**Results:** Booster uptake was 68% among those aged ≥60 and 30% among those aged 18-59 years with a medical risk factor for severe disease. For both target groups the most important determinant for booster uptake was age (15% in 18-29 years to 72% in 80+ years). Voting proportions for progressive liberal political parties ranked second in the random forest analysis in both groups, with an increasing proportion of votes associated with higher uptake. In the 60+ group, household type ranked third, with highest vaccine uptake among married couples without children (72%) and the lowest uptake among unmarried couples with children (47%). In the medical risk group, migration status ranked third. Migrants with two parents born abroad had the lowest uptake (18%), whereas migrants with both parents born in the Netherlands had the highest uptake (35%).

**Conclusion:** Among individuals who had received at least one prior COVID-19 vaccination, the autumn 2022 COVID-19 booster uptake was 68% in people ≥60 years and 30% in in the medical risk group aged 18-59 years. The most important determinant of booster uptake was age, followed by political preference and household type (60+ group) or migration status (medical risk group). Uptake varied considerably among subgroups in both target groups. Further research should be aimed at understanding the drivers and barriers of vaccine uptake among the subgroups with notably low uptake.

## Introduction

With the introduction of COVID-19 vaccines in January 2021, the cornerstone of containing the COVID-19 pandemic was laid. Vaccine effectiveness against severe disease and mortality of the primary vaccination series was high (1). However, because of waning of vaccine effectiveness and the emergence of new SARS-CoV-2 variants, several booster vaccination campaigns were implemented with the aim of restoring protection against severe disease. In the Netherlands, the first COVID-19 vaccination campaign for the entire population started in January 2021, followed by a first booster campaign that started in November 2021. In Spring 2022, another booster vaccination was offered to clinically vulnerable people (2). A booster with a bivalent COVID-19 vaccine was offered from 19 September 2022 onwards in the so-called autumn booster campaign. This autumn booster was offered to the Dutch population aged 12 years and older, and targeted especially clinically vulnerable individuals (medical risk group) and individuals aged 60 years and older (3). People with a medical risk and elderly people are at higher risk of developing severe disease after SARS-CoV-2 infection (4) and a high vaccination uptake in this group is thought to be essential to avoid excess morbidity and mortality (5, 6, 7).

However, previous research showed that the autumn 2022 booster uptake in the medical risk group aged 18-59 years in the Netherlands was low (28%) (8). Therefore, further research into the determinants of vaccination among this group was required. In two studies conducted in Australia and the United States among clinically vulnerable people, being unvaccinated was associated with age, sex, education level, income and ethnicity (9, 10).

However, sample sizes were small and the same determinants may not be applicable in the Netherlands. Consequently, the aim of the current study was to investigate vaccine uptake during the autumn 2022 booster campaign among the population subgroups at risk for severe COVID-19 that were specifically targeted by this campaign: people with a medical risk aged 18-59 years and individuals ≥60 years, who had received at least one previous COVID-19 vaccination. We calculate the booster uptake in each of these populations and analyze the importance of potential determinants of the booster uptake.

## Methods

### Study population

The study population consisted of the Dutch population aged 18 years and older, registered both in the Personal Records Database and the COVID-19-vaccination Information and Monitoring System (CIMS), maintained by the Dutch National Institute for Public Health and the Environment (RIVM), and with at least one COVID-19 vaccination before the autumn 2022 booster round. Determinants of vaccination were studied in people aged ≥60 years (including individuals with and without medical risk) and in the medical risk group also in people aged 18-59 years. These populations were determined on 18 September 2022, the day before the start of the autumn 2022 booster campaign. Medical risk group status was based on Dutch national healthcare registry data which contained information from a claims database of outpatient specialist care utilization and data on medication at ATC-4 code level, complemented with data on long-term care utilization. Medical risk was defined by the method described by Pijpers et al. (11). In short, individuals with a high risk are those with one or more comorbidities that are associated with a high risk for severe outcomes of SARS-CoV-2 infection (12). The intermediate risk group consists of people with an indication for the annual influenza vaccination (12). Individuals living in a nursing home and/or having an intellectual disability were also prioritized in the vaccination campaign and were classified as ‘other’.

### Vaccination data

The autumn 2022 booster campaign started on 19 September 2022. An autumn 2022 booster dose was defined as a COVID-19 vaccination administered since 19 September 2022 among individuals who had at least one registered COVID-vaccination (since primary vaccination consisted of at least one vaccination, e.g. in case of earlier positive test).

Vaccination data were obtained from CIMS (extraction date: 7 March 2023). Only data of individuals who gave informed consent to register their vaccination data are included in the CIMS database. For the primary vaccination series, informed consent was given by 93% of all vaccinated individuals (13). If vaccinated individuals did not give informed consent for the primary vaccination to be registered, they could not be distinguished from unvaccinated individuals and are thus not included in the study. Of those who received an autumn 2022 booster, 99.2% provided informed consent to be registered in CIMS (8). People who did not provide informed consent for the autumn 2022 booster to be registered in CIMS were regarded as unvaccinated.

### Determinants of vaccination

Within the remote access environment of Statistics Netherlands (CBS), population and vaccination data were linked at individual level (using a unique identifier) to other national databases containing information on the following potential determinants: age, sex, education level, origin (consists of two variables “country of origin” and “migration status”), socioeconomic position, personal income, household type, household car ownership, employment sector, urbanization level, and medical risk group. Voting proportions for different political movements and place of residence (expressed as X-Y coordinates of 6-digit postal codes) were linked at neighborhood level. An elaborate description of these determinants can be found in Supplemental Table 1.

### Statistical analyses

The same statistical methods were used as in the study by Pijpers et al. (11). Briefly, a random forest (RF) analysis was performed for each of the two target populations, to rank sociodemographic determinants according to their importance in the prediction of COVID-19 autumn booster uptake. The results of the RF analyses were visualized by ranking determinants based on the increase in probability of misclassification (PMC) that results from replacing the values of a particular determinant by random values. Furthermore, descriptive analyses were performed to interpret the direction of the associations of the various determinants. To avoid small subgroups producing a distorted image of the general situation within a target population, frequencies below 100 were excluded in the heatmaps. In the figures and descriptive analyses, age was divided in categories of ten years. For the figures of the 60+ group, only the categories ‘60-69’ and ‘70+’ were used, since little difference in uptake of the booster vaccination was found among the higher age groups. In the RF analysis of the 60+ group, the determinants ‘employment sector’ and ‘education level’ could not be included because of among those employment sector was largely not applicable and for educational level in this age group the proportion of missing data is large (14). All analyses were done using RStudio, version 4.1.3.

### Ethics statement

The Centre for Clinical Expertise at the RIVM assessed the study and stated that it was exempt from the law for medical research involving human subjects (WMO). In line with publication guidelines of Statistics Netherlands, all percentages and numbers are rounded to the nearest ten (15).

## Results

### People aged ≥60 years

The Dutch population aged ≥60 years that received at least one previous COVID-19 vaccination comprised of 4,023,620 individuals. By 7 March 2023, 68% of this population had received an autumn 2022 booster. Uptake was approximately the same among individuals aged 60 and above with a medical risk (68%) and without (67%). Vaccine uptake per determinant and across age groups is provided in Supplemental Tables 2 and 3.

### Ranking of determinants

Figure 1 shows the ranking of the determinants among those aged ≥60 years, based on the results of the RF analysis. The performance characteristics of the RF are a PMC of 39%, a sensitivity of 61% and a specificity of 60%.

**Figure 1.**
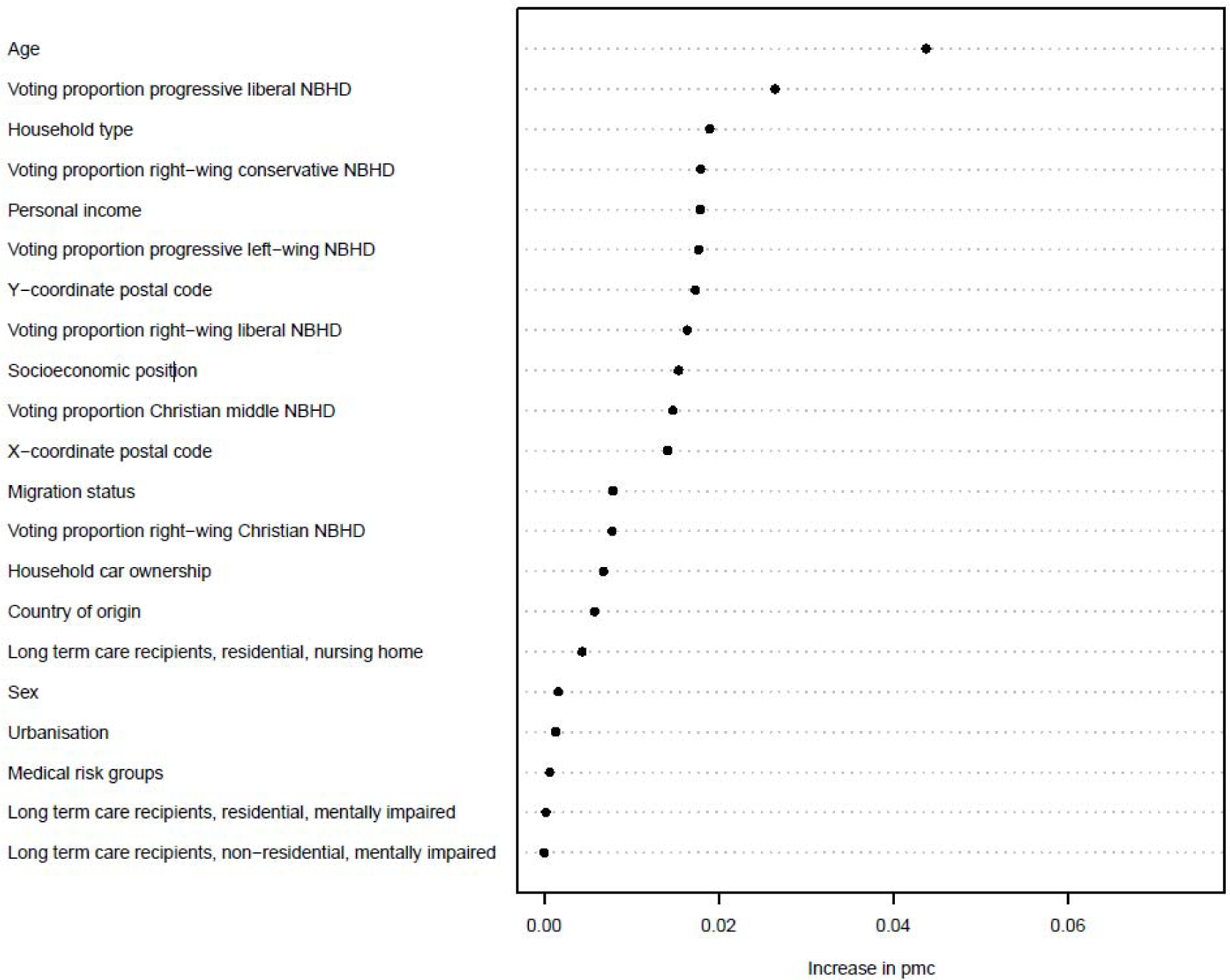
Ranking of variable importance for predicting the 2022 COVID-19 autumn 2022 booster uptake in people aged ≥60 years of age. *NBHD* = neighborhood.

### Age

By far the most important determinant was age (Supplemental Table 2). Autumn 2022 booster uptake increased with age, except for people aged 80+ years who had a slightly lower uptake than people aged 70-79 years.

### Voting proportions for political parties

Voting proportions for progressive liberal parties ranked second in the RF, while voting proportions for other political movements ranked lower. Only voting proportions for right-wing Christian parties (ranked 13^th^) contributed much less than voting proportions for the other political movements to the increase in PMC. Figure 2 shows the autumn 2022 booster uptake per age group by increasing proportions of votes in neighborhoods for each political movement. Autumn 2022 booster uptake increased with higher proportions of votes for progressive liberal and right-wing liberal parties. For right-wing conservative and right-wing Christian parties, the opposite was true with lower uptake with increasing voting proportions. For Christian middle political parties, uptake did not vary as much with increasing voting proportions. With respect to progressive left-wing parties, uptake was lowest for people in neighborhoods with either a very low or a very high proportion of votes. Overall, uptake was higher in older age groups (Supplemental Table 2).

**Figure 2.**
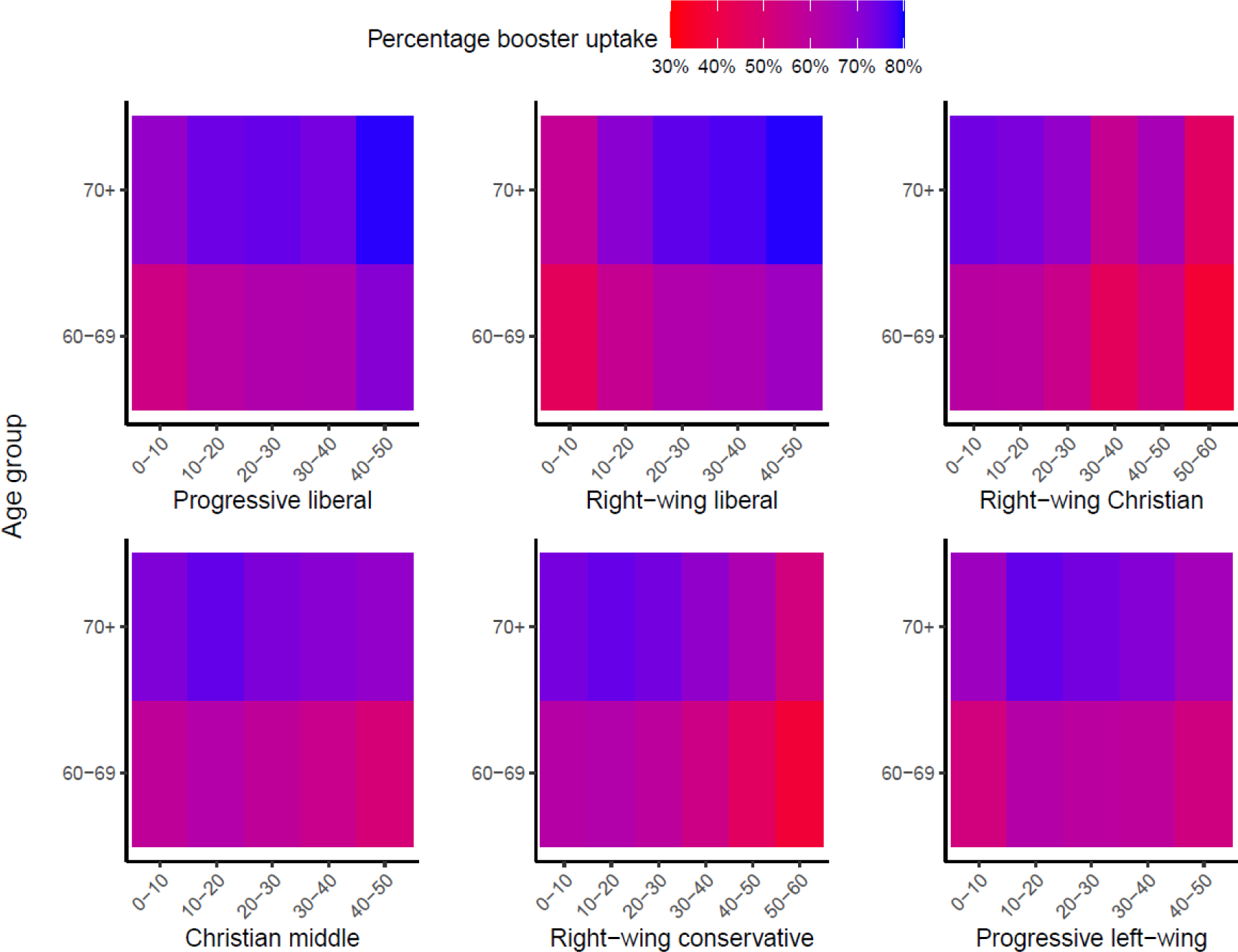
Autumn 2022 booster uptake in the 60+ population, by voting proportions (%) for political movements in the 2021 elections at neighborhood level, per age group. *Color gradient is adjusted based on the minimum and maximum uptake for the specific determinants in this figure*.

### Household type

Household type ranked 3^rd^ in the RF for those aged ≥60 years. Autumn 2022 booster uptake was highest among married couples without children (72%) and lowest in single-parent households (48%) and unmarried couples with children (47%) (Figure 3 and Supplemental Table 2).

**Figure 3.**
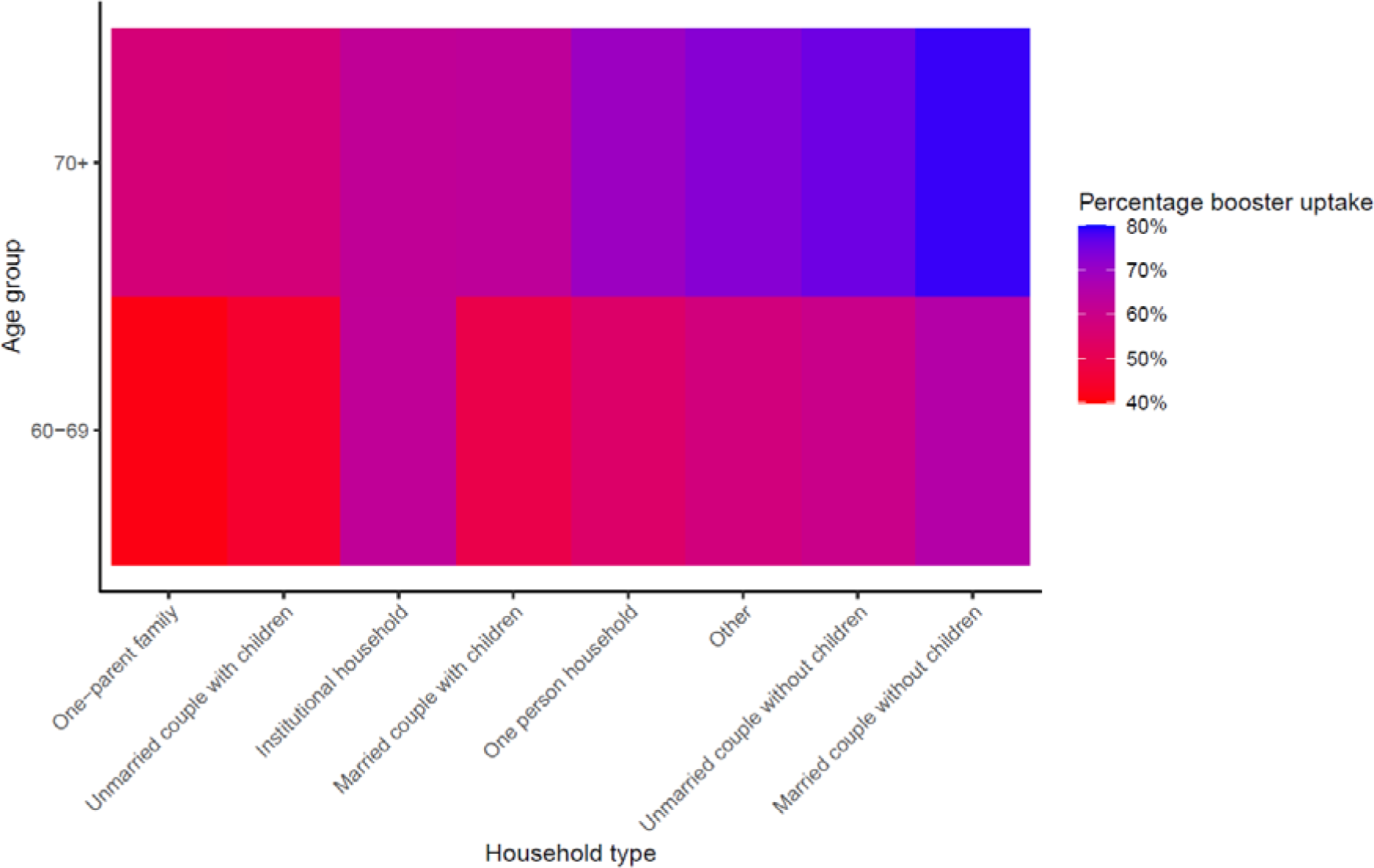
Autumn 2022 booster uptake in the 60+ population, by household type and age group. *Color gradient is adjusted based on the minimum and maximum uptake for the specific determinants in this figure. With/without children denotes children living in the same house*.

### Personal economic situation

The variables personal income and socioeconomic position ranked 5^th^ and 9^th^ in the RF for people ≥60 years. Autumn 2022 booster uptake was lowest (56%) for individuals with the lowest income group and increased with increasing income to the highest uptake (71%) for individuals in the highest income decile. With respect to socioeconomic position, uptake was lowest for individuals receiving social assistance benefits (28%) and highest amongst pensioners (72%) (Figure 4 and Supplemental Table 2).

**Figure 4.**
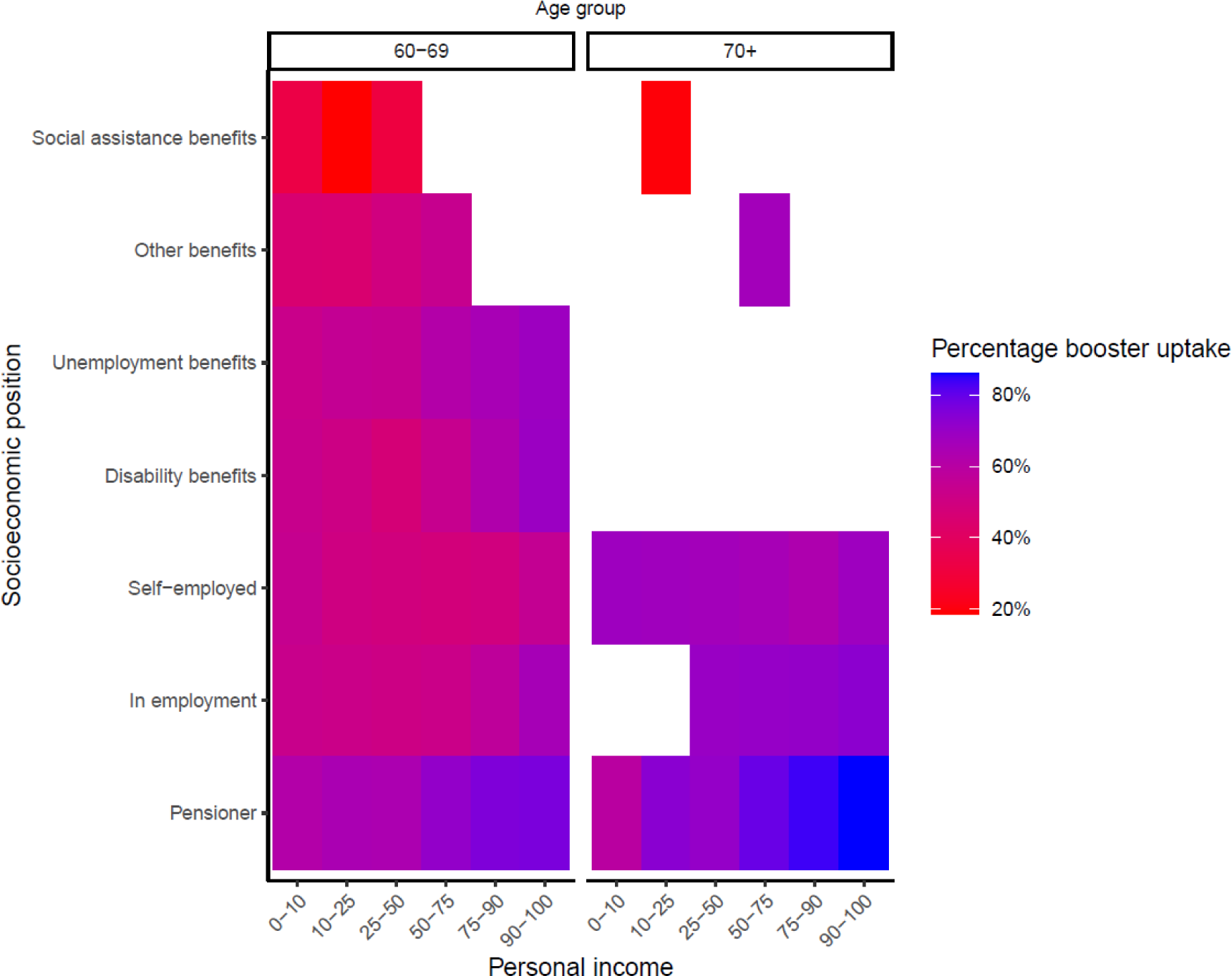
Autumn 2022 booster uptake in the 60+ population, by socioeconomic position and personal income (percentage intervals), per age group. *Color gradient is adjusted based on the minimum and maximum uptake for the specific determinants in this figure*.

### X-Y coordinates of the postal code

A lower booster uptake among those aged ≥60 years was mainly concentrated in the Province of Friesland, the eastern part of the Province of Groningen and in areas where a relatively high proportion of the population is of orthodox reformed denomination (Supplemental Figure 1).

### People with a medical risk aged 18-59 years

At the extraction date, the medical risk group aged 18-59 years comprised of 1,559,600 individuals (16% of the entire Dutch population of this age group). Of these, 1,340,400 individuals (86%) had received at least one previous COVID-vaccination. Of those, 30% received an autumn 2022 booster (28% of the group with intermediate risk and 34% of the high risk group).

Figure 5 shows the ranking of the determinants, based on the results of the RF analysis. Performance characteristics of the RF are a PMC of 34%, a sensitivity of 66% and a specificity of 65%.

**Figure 5.**
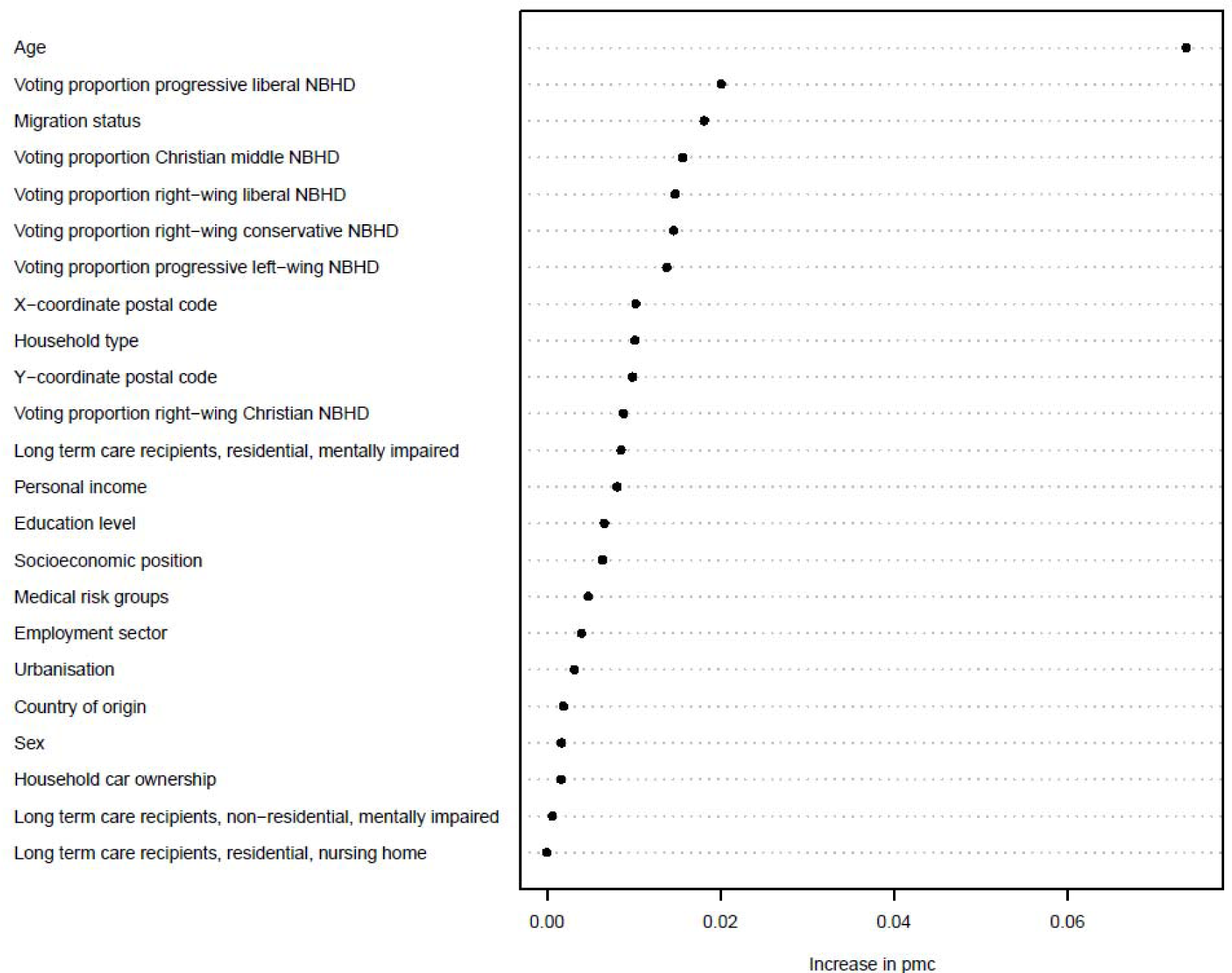
Ranking of variable importance for predicting COVID-19 autumn 2022 booster uptake in people with a medical risk aged 18-59 years.

### Age

In this medical risk population, age was also the most important determinant. Autumn 2022 booster uptake increased with increasing age, for each determinant, as shown in Supplemental Table 3.

### Voting proportions political parties

Furthermore, voting proportions for political movements in the 2021 national elections were relatively important determinants, except for the right-wing Christian movement. For progressive liberal and right-wing liberal parties, autumn 2022 booster uptake was higher with increasing proportions of votes. For the right-wing conservative movement, the opposite was true: autumn 2022 booster uptake was lower with increasing proportions of votes. Regarding the progressive left-wing movement, uptake was lowest for people in neighborhoods with either a very low or a very high proportion of votes (Figure 6). Overall, these trends were most pronounced for the higher age groups (40-49 and 50-59 years). Compared to the results in the 60+ group, a similar booster uptake pattern was seen for the progressive liberal, right-wing liberal, right-wing conservative and progressive left-wing movements, although booster uptake was generally lower in the medical risk group aged 18-59 years, especially in the younger age categories (18-29 years and 30-39 years).

**Figure 6.**
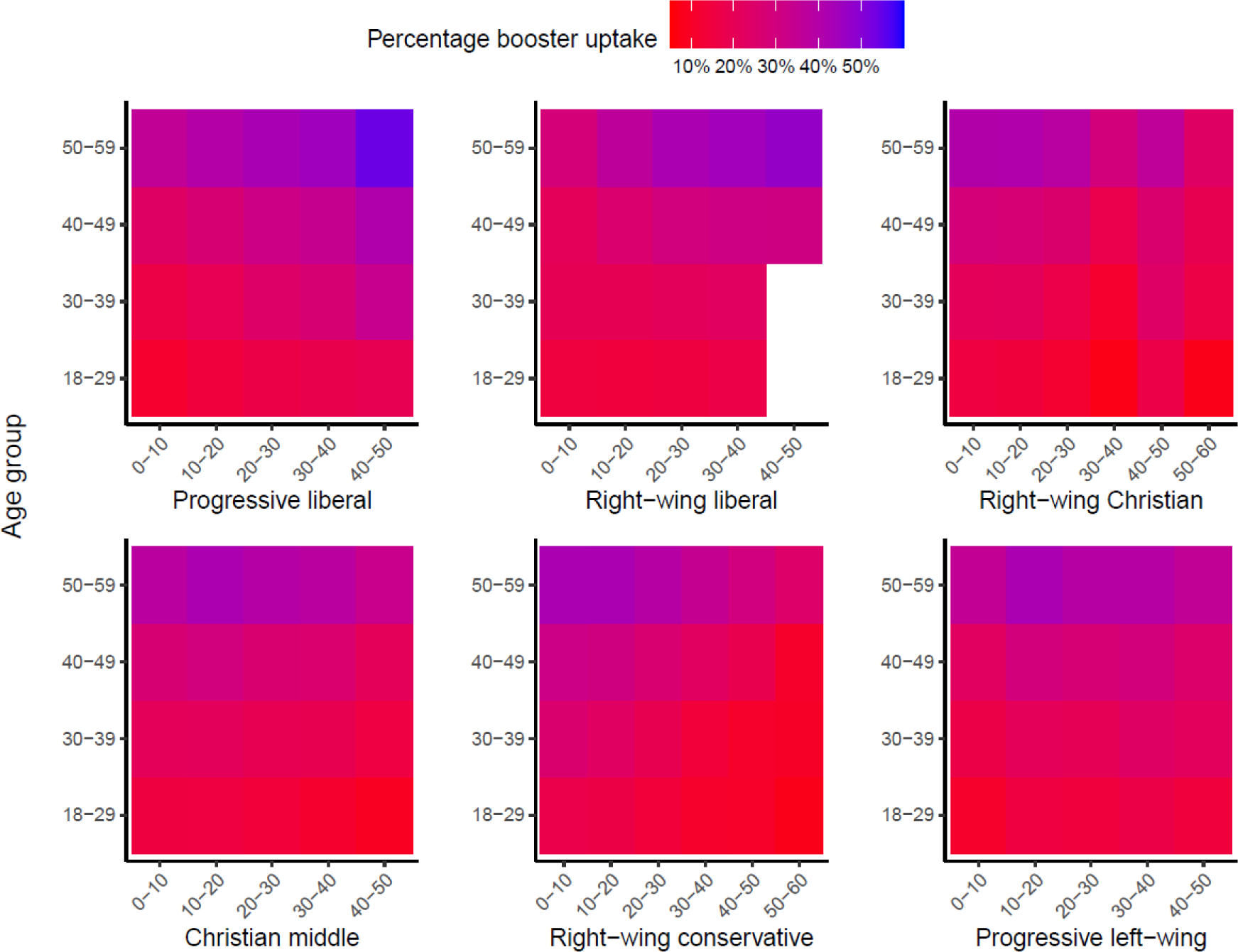
Autumn 2022 booster uptake for the medical risk group, by voting proportions (%) for political movements in the 2021 elections at neighborhood level, by age group. *Color gradient is adjusted based on the minimum and maximum uptake for the specific determinants in this figure*.

### Migration status

Migration status ranked third in the medical risk group aged 18-59 years (Figure 7). Migrants with two parents born in the Netherlands had the highest booster uptake (35%), whereas migrants and children of migrants with two parents born abroad had the lowest booster uptake (18% and 17%, respectively). In contrast to migration status, country of origin ranked low in the RF analysis despite considerable variation between countries/regions of origin. Autumn 2022 booster uptake was lowest in people born in Morocco (5%) and Turkey (7%) and highest in people born in Indonesia, the Netherlands and Other EU countries (38%, 33% and 33%, respectively) (Figure 8 and Supplemental Table 3).

**Figure 7.**
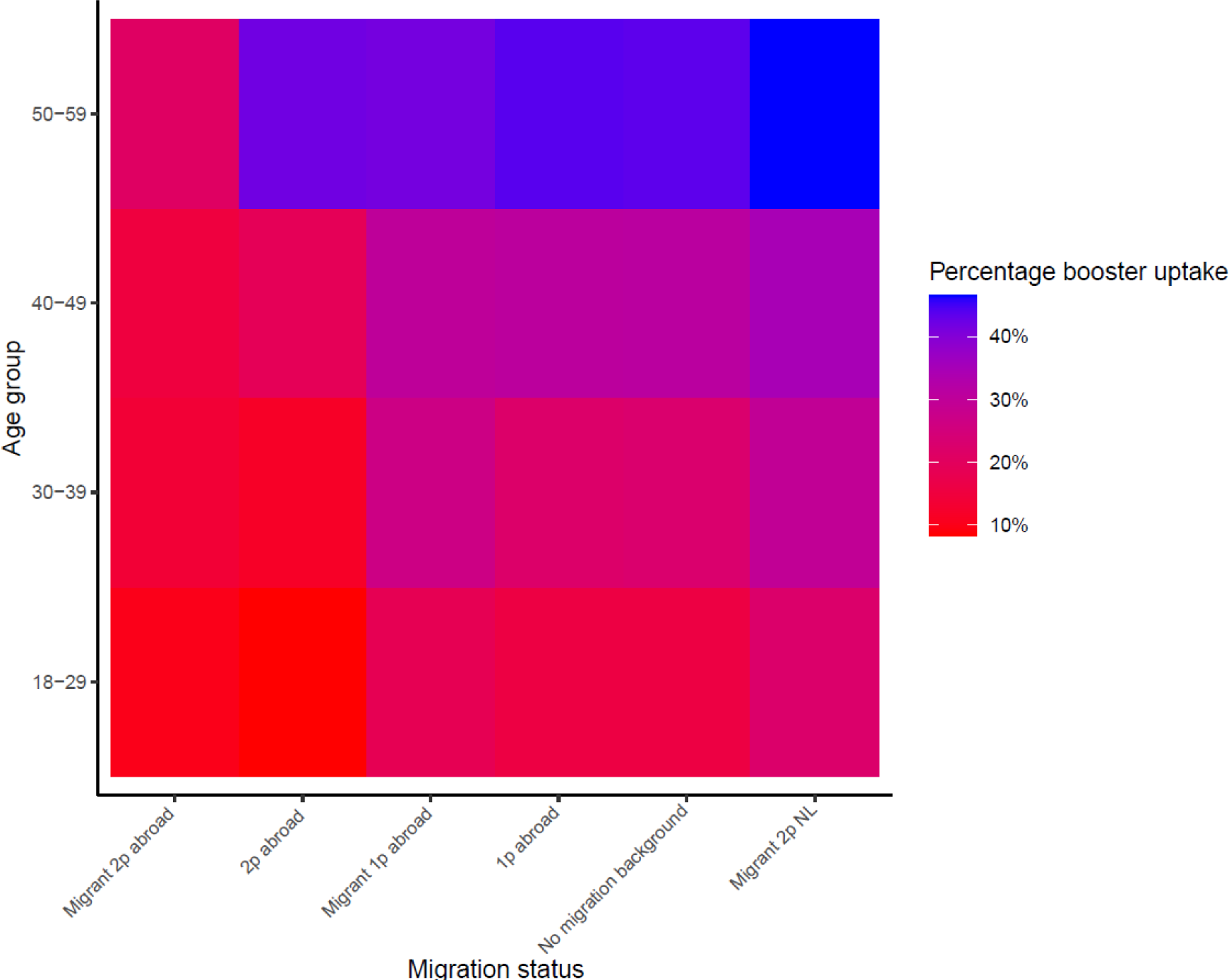
Autumn 2022 booster uptake for the medical risk group by migration status and age group. Abbreviations: *2p* two parents, *1p* one parent, *NL* The Netherlands. *Color gradient is adjusted based on the minimum and maximum uptake for the specific determinants in this figure*.

**Figure 8.**
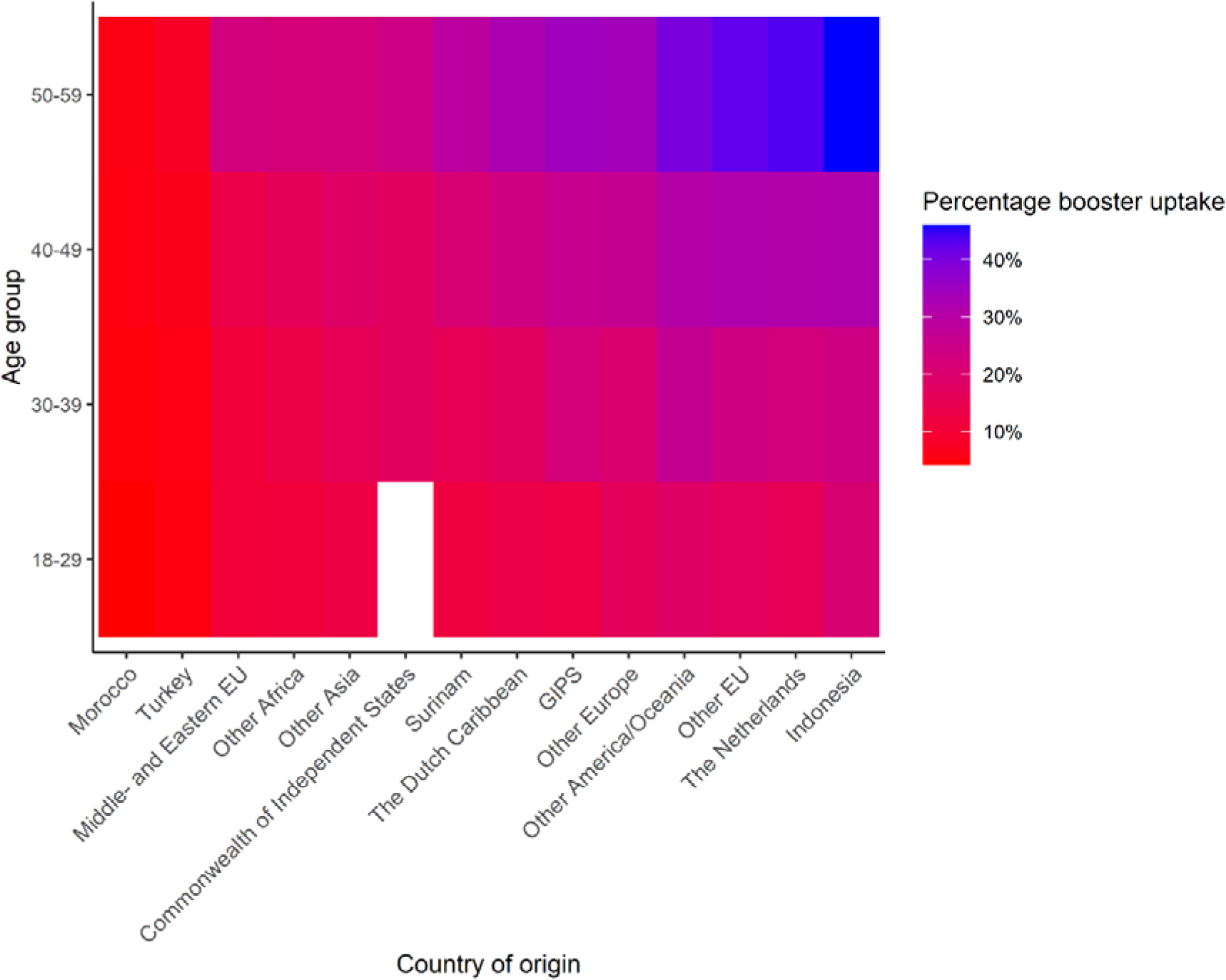
Autumn 2022 booster uptake for the medical risk group by country of origin, per age group. Abbreviations: *EU* European Union, *GIPS* Greece, Italy, Portugal, Spain. *Color gradient is adjusted based on the minimum and maximum uptake for the specific determinants in this figure*.

## Discussion

Of the 90% of people aged ≥60 years in the Netherlands who received the primary vaccination against COVID-19 (11), 68% opted to receive the autumn 2022 booster. In the previously vaccinated medical risk group aged 18-59 years, the autumn 2022 booster uptake was only 30%. In both target groups, age was the most important determinant of the autumn 2022 booster uptake. Our bivariate analyses showed that in general, across all determinants, uptake steeply increased with age, although the increase flattened above 69 years. In addition to higher age, a high voting proportion for progressive liberal parties within the neighborhood ranked for both target groups as the second most important determinant for uptake. In the 60+ group, being married without children living in the same household, X-Y coordinates for postal code and a high personal income were also relatively important determinants for booster uptake. In the medical risk group aged 18-59 years, migration status ranked third.

The much lower uptake of the autumn 2022 booster among people aged ≥60 years compared to the primary COVID-19 vaccination series was not surprising, as uptake was already declining with the first booster in Autumn 2021 (16). It is also consistent with findings from many other countries, where a decline in COVID-19 vaccine uptake is seen in each following vaccination round (17, 18, 19, 20, 21). Moreover, the autumn 2022 booster uptake in the entire population aged ≥60 years (including people without a previous vaccination) was similar to seasonal influenza uptake in this age group: 67.5% versus 65% (22, 23).

However, the autumn 2022 booster uptake of only 30% in the medical risk group aged 18-59 years was lower than expected, given their increased risk of severe outcomes (e.g. hospitalization and death) of COVID-19 (5) and considering that they were specifically targeted by this vaccination campaign (they received a personal invitation from their general practitioner). Even though the autumn 2022 booster uptake in the medical risk group was low, it was higher compared to that among same aged individuals without a medical risk (17%; data not shown). Furthermore, it was similar to the 2021 influenza vaccine uptake in people aged 18-59 years with a medical risk (33%) (22). Although the 2022 autumn COVID-19 booster uptake in the medical risk population aged 18-59 was calculated amongst those who had received a at least one previous COVID-19 vaccination, the uptake is roughly comparable. Unfortunately, determinants of influenza uptake cannot be assessed on a national level, since there is no nationwide influenza vaccination register.

A possible explanation for the low(er) autumn 2022 uptake is that this vaccination round was during the Omicron era. In this phase of the pandemic, COVID-19 was less severe than earlier SARS-CoV-2 variants, with a shorter period of illness and reduced probability of hospital admission and mortality (24, 25). There were no more behavioral restrictions and there was generally less concern about COVID-19 infections in the Dutch society and elsewhere. Also, many people had had an infection or were vaccinated at the time (seroprevalence was >95%) (26). Therefore, the perceived necessity of getting another COVID-19 booster may have been low.

Other studies also report that a lower level of perceived susceptibility to severe COVID-19 infection and having gone through an infection are reasons for not being vaccinated with the booster dose among individuals who were previously vaccinated (27, 28, 29).

In our study, important determinants for the autumn 2022 booster uptake were age, political preference, household type, income and migration status. Among the 60+ population, age was most important, followed by political preference, household type, income and X-Y coordinates. A similar analysis focused on the *primary vaccination* uptake among people aged ≥60 years in the Netherlands found that the most important determinants were political preference, X-Y coordinates and personal income, whereas age and household type ranked low (11). These discrepancies may be (partially) due to the fact that uptake of the primary series was determined relative to the entire Dutch population of 18 years and older whilst the autumn 2022 booster uptake was determined relative to the population that had received at least one prior vaccination.

Our results are in concordance with findings from other studies investigating uptake of doses other than the first. The trends we observed in our bivariate analyses, with a lower uptake associated with being younger, unmarried, living in urban areas, belonging to ethnic minority groups and having a lower education, were also found by Wang et al. and Tessier et al. (20, 30). Among the people aged 18-59 years with a medical risk, migration status was an important determinant for autumn 2022 booster uptake. Two studies among people with chronic conditions also found that ethnicity was associated with vaccination status (9, 10).

Although migration status ranked high in the medical risk group aged 18-59 years, country of origin ranked only 19^th^. Since these determinants are closely related, this is a somewhat unexpected result. The RF analysis cannot explain how the different determinants interact to constitute a prediction of the importance of determinants for vaccination uptake. However, a plausible explanation is that within the interplay of the other determinants age, political preference, personal income, education level, household type etc., it is more important whether a person has a migration background than what the exact country of origin of the person itself or his/her parents is. The known barriers to vaccine uptake in migrants, such as language and communication barriers, cultural or religious barriers, practical barriers and distrust of health system or authorities (31), are applicable to migrants from various countries of origin.

In the bivariate analyses, some subgroups with a low autumn 2022 booster uptake were identified. Subgroups (of both target populations) with a notably low uptake were single-parent households, people living in highly urbanized areas, people receiving social assistance benefits, people from the lowest income groups, migrants with two parents born abroad and people from neighborhoods with a high voting percentage for right-wing Christian or right-wing conservative political movements. Further analyses should be conducted to gather information on the specific reasons for non-vaccination within these groups.

The major strength of this study is the use of nation-wide data, enhancing the generalizability of our findings. In addition, most information was available at individual level, except for voting proportions for political movements, which were available at neighborhood level. Moreover, in contrast to some studies about vaccine hesitancy we were able to study autumn booster vaccine uptake directly rather than intention to vaccinate. This type of research based on national registry data is very important to inform vaccination strategies. Given the important inequity (32) we found, this type of research is a priority for other vaccination programs in the Netherlands, such as infant vaccination and for example influenza and pneumococcal vaccination in adults. A prerequisite for it is, however, the existence of a nationwide register.

There are several limitations to this study that should be kept in mind when considering the results. Firstly, the most recent data available on medical conditions was from 2020. This may have led to considerable misclassification of individuals if they have received a diagnosis since then. Also, for some conditions the data used to identify whether an individual belonged to a medical risk group were not very specific. For instance, all people receiving medication for lung conditions were classified as having a lung condition and, therefore, part of the medical risk group, even though they may not need/use the medication. Furthermore, there is a small percentage (0.8%) of people who did not provide consent for their autumn booster vaccination data to be registered. They were considered unvaccinated in the analyses, which may have biased the results to some extent. The fact that the population was extracted at one point in time may have added to this bias, because we did not take into account people who became 60 years between September and March 2022, nor people who died during this period. Also, data on previous infections were not available, whereas a recent infection may have been a reason not to get the autumn 2022 booster.

Also, people receiving the second dose of the primary vaccination may have been falsely classified as having an autumn 2022 booster, since booster uptake was based on vaccination date. However, the number of people aged ≥18 years getting the primary series is expected to be very small. In addition, our study population consisted only of individuals who had received a primary vaccination *and* gave informed consent for this data to be shared. We know that older people are more willing to provide informed consent (personal communication S. McDonald), but for other characteristics (such as medical risk group status), this information is unknown. Moreover, data on other relevant determinants of vaccination such as religion, personal motivation and self-efficacy and previous COVID-19 infections were not available. This is a likely explanation for the relatively low predictive value and validity of the entire set of determinants we were able to include in the analyses: the probability of misclassification for both target groups was over 30% (39% 60+ group and 34% medical risk group) and sensitivity and specificity were around 60% for both groups. It is also reflected by the high ranking of variables at neighborhood level (political preference and in the 60+ group also X-Y coordinates), indicating that there are unmeasured variables that contribute to vaccine uptake.

## Conclusion

In conclusion, one target group at risk for severe disease was much better reached with the autumn 2022 booster (68% uptake in people aged ≥60 years) than the other target group (30% uptake in people with a medical risk aged 18-59 years). We showed that within these groups, age was the most important determinant for autumn 2022 booster uptake. Other important determinants were political preference, personal income, household type, X-Y coordinates and migration status. The findings of this study can be used in future vaccination strategies, especially when targeting clinically vulnerable and elderly people. Further research should be aimed at understanding the drivers and barriers of vaccine uptake among the subgroups with notably low uptake.

## Data Availability

All data is available within CBS Microdata and can be made available under strict conditions.

## Notes

### Competing Interest Statement

The authors have declared no competing interest.

### Funding Statement

This study was funded by the Dutch Ministry of Health, Welfare and Sports

### Author Declarations

The Centre for Clinical Expertise at RIVM assessed the research proposal (EPI–613). They verified whether the work complies with the specific conditions as stated in the law for medical research involving human subjects (WMO). They are of the opinion that the research does not fulfil one or both of these conditions and therefore it is excluded for further approval by the ethical research committee.

