## Supplemental information for "Determinants of COVID-19 booster uptake in the Netherlands, autumn 2022: how well were those at risk for severe disease reached?"

**Supplemental Figure 1.** Autumn 2022 booster uptake for the population aged ≥60 years per neighborhood.

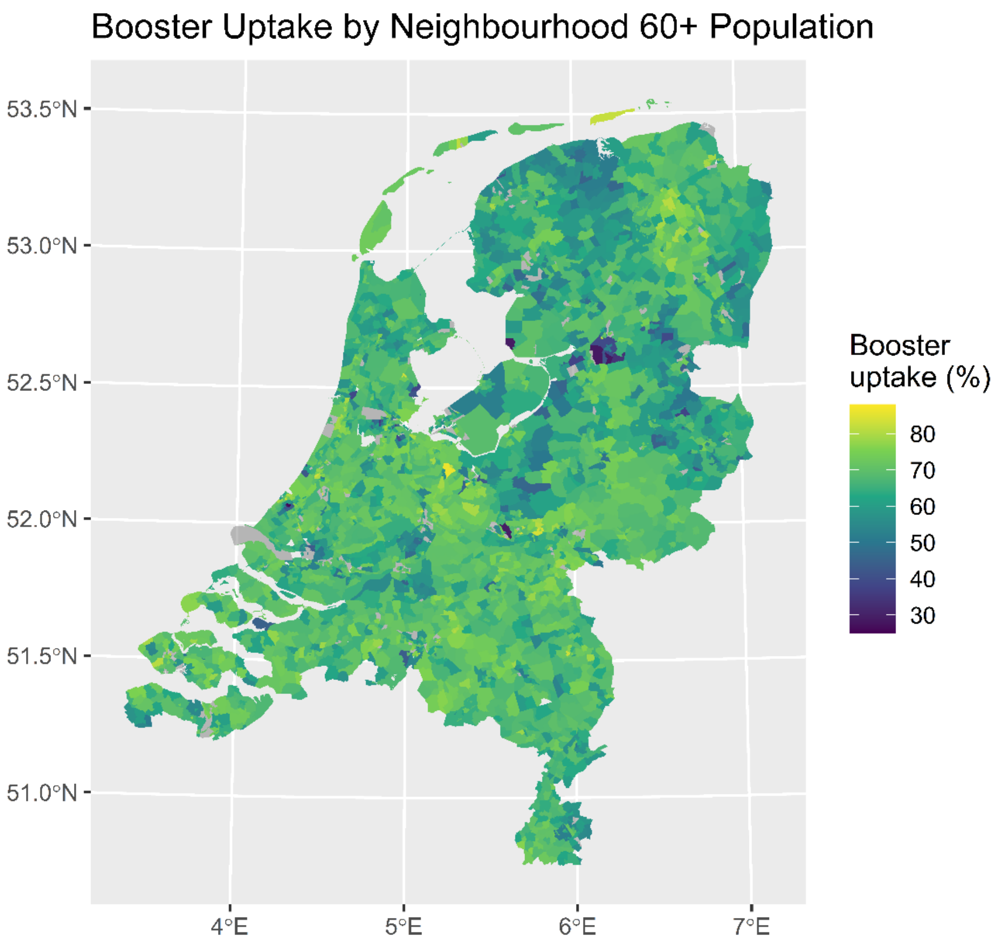

Note: the grey areas indicate neighborhoods with frequencies <10 and were therefore excluded.

**Supplemental table 1: Description of determinants**

| Variable | Levels | Reference date (dd-mm-yyyy) | Database |
|---|---|---|---|
| ***Individual level*** | | | |
| Vaccine uptake | At least one COVID-19 vaccination registered in CIMS; 1 = yes 0 = no[a] | 19-09-2022 – 06-03-2023[b] | CIMS |
| Age | Continuous[c] | 18-09-2022 | CBS |
| Sex | 0 = male; 1 = female | 18-09-2022 | CBS |
| Education level | Primary education; prevocational secondary education-basic vocational programme (VMBO-b/k), lower secondary vocational training and assistant's training (MBO-1); prevocational secondary education – theoretical and vocational programme (VMBO-g/t), the first three years of senior general secondary education (HAVO) and pre-university secondary education (VWO); basic vocational training (MBO-2) and vocational training (MBO-3); middle management and specialist education (MBO-4); upper secondary education (HAVO/VWO); Hbo-, wo-bachelor; Hbo-, wo-master, doctor; Unknown | 18-09-2022 | CBS |
| Country of origin[d] | The Netherlands; Turkey; Morocco; Surinam; The Dutch Caribbean; Indonesia; Other Africa; Other Asia; Other America/Oceania; Middle and Eastern European countries within the EU; GIPS countries (Greece, Italy, Portugal, Spain); Former or associated member states of the Commonwealth of Independent States, Other countries of the EU; Other European countries; Unknown | 18-09-2022 | CBS |
| Migration status[,f] | Netherlands; Born in the Netherlands with one parent born abroad; Born in the Netherlands with two parents born abroad; Born abroad with one parent born abroad; Born abroad with two parents born abroad; Born abroad with two parents born in the Netherlands; Unknown | 2022 | CBS |
| Socioeconomic position | In employment; Self-employed; Unemployment benefits (WW); Social assistance benefit; Other benefits; Disability benefit; Pensioner; Student; Other/Unknown | 2022 | CBS |
| Personal income | Continuous (percentiles)[g] | 2022 | CBS |
| Household type | One person household; Unmarried couple without children; Married couple without children; Unmarried couple with children; Married couple with children; One-parent family; Other households; Institutional household; Other. *With/without children denotes children living in the same house.* | 2022 | CBS |
| Household car ownership | 1 = yes; 0 = no | 2022 | CBS |

| | | | |
|---|---|---|---|
| Employment sector | Agriculture, Forestry and fishery; Mining and quarrying; Industry; Electricity supply; Water supply, sewerage and waste management; Construction; Wholesale and retail trade; Transportation and storage; Accommodation and food service activities; Information and communication; Financial services; Real estate activities; Professional scientific and technical activities; Administrative and support service activities; Public administration and defence; Education; Human health and social work activities; Arts, entertainment and recreation; Other service activities; Activities of household as employer, undifferentiated goods- and service producing activities of households for own use; Activities of extraterritorial organisations and bodies; Other/unemployed/unknown | 2022 | CBS |
| Urbanisation level | Not urbanised; Hardly urbanised; Moderately urbanised; Strongly urbanised; Extremely urbanised; Unknown | 2022 | CBS |
| X-coordinate postal code | Numeric | 2022 | CBS |
| Y-coordinate postal code | Numeric | 2022 | CBS |
| Medical risk groups[h] | High medical risk; Intermediate medical risk; Low medical risk | 2020[i] | CBS |
| Long term care recipients, residential, nursing home | 1 = yes; 0 = no | 2022 | CBS |
| Long term care recipients, residential, mentally impaired | 1 = yes; 0 = no | 2022 | CBS |
| Long term care recipients, non-residential, mentally impaired | 1 = yes; 0 = no | 2022 | CBS |
| ***Neighbourhood (NBHD) level*** | | | |
| Voting proportions for political movement[h] | | 2021 | Open State Foundation |
| Right-wing liberal (VVD) | Percentage | | |
| Progressive liberal (D66, Volt) | Percentage | | |
| Christian middle (CDA, CU) | Percentage | | |
| Right-wing Christian (SGP) | Percentage | | |
| Progressive left-wing (GL, PvdA, PvdD, SP, DENK) | Percentage | | |

| | |
|---|---|
| Right-wing conservative (PVV, FvD, JA21) | Percentage |
| Other parties | Percentage |

*Abbreviations: CBS Centraal Bureau voor de Statistiek (Statistics Netherlands), CDA* Christian Democratic Appeal, *CU* Christian Union, *D66* Democrats 66, *FvD* Forum for Democracy, *GL* Green Left, *JA21* Right Answer 2021, *PvdA* Labour party, *PvdD* Party for the Animals, *PVV* Party for Freedom, *SP* Socialist Party, *Volt* Volt Netherlands, *SGP* Reformed Political Party, *VVD* People's Party for Freedom and Democracy

a) 0 = did not receive vaccination or did not provide informed consent for data to be registered in CIMS

b) The extraction date was 07-03-2023

c) Age was estimated as 2021 minus year of birth

d) Both country of origin and migration status were included in the analysis, based on the new classification (level 3) of population by origin by CBS. More information on the CBS 2022 Classification of population by origin can be found at: New classification of population by origin (cbs.nl)

e) If a person was born in the Netherlands and their mother was born abroad, country of origin was defined as the mother's country of birth. If only the father was born abroad, their father's country of birth was the person's country of origin.

f) Percentiles are calculated by CBS Microdata based on personal income data including the entire Dutch population in 2022

g) Approximated based on healthcare utilisation and medication prescription data

h) In case of rare medical conditions, data from 2016-2020 were included.

i) Based on information from the National Elections in March 2021 for political parties with at least 2 seats. Data available from the Open State Foundation URL: Kiesraad | Data overheid.

**Supplemental Table 2. Autumn booster uptake per determinant and age group for individuals aged ≥60 years.**

| | N | Autumn booster uptake (N, %) |
|---|---|---|
| **Age** | | |
| 60-69 | 1.923.200 | 1.157.650 (60) |
| 70-79 | 1.448.490 | 1.084.820 (75) |
| 80+ | 651.940 | 468.890 (72) |
| **Sex** | | |
| Male | | |
| 60-69 | 957.520 | 573.070 (60) |
| 70-79 | 694.610 | 530.210 (76) |
| 80+ | 255.840 | 190.280 (75) |
| Female | | |
| 60-69 | 965.680 | 584.580 (61) |
| 70-79 | 753.870 | 554.610 (74) |
| 80+ | 396.100 | 278.070 (70) |
| **Household car ownership** | | |
| Yes | 3.245.030 | 2.249.380 (69) |
| 60-69 | 1.662.070 | 1.028.040 (62) |
| 70-79 | 1.202.070 | 930.520 (77) |
| 80+ | 380.880 | 290.810 (76) |
| No | 778.600 | 461.980 (59) |
| 60-69 | 261.130 | 129.600 (50) |
| 70-79 | 246.420 | 154.290 (63) |
| 80+ | 271.060 | 178.080 (66) |
| **Country of origin** | | |
| The Netherlands | 3.434.940 | 2.384.400 (69) |
| 60-69 | 1.615.470 | 1.011.060 (63) |
| 70-79 | 1.253.190 | 959.850 (77) |
| 80+ | 566.280 | 413.500 (73) |
| Turkey | 27.190 | 3.990 (15) |
| 60-69 | 15.740 | 1.950 (12) |
| 70-79 | 8.520 | 1.460 (17) |
| 80+ | 2.930 | 570 (19) |
| Morocco | 26.550 | 2.180 (8) |
| 60-69 | 14.520 | 1.000 (7) |
| 70-79 | 8.620 | 900 (10) |
| 80+ | 3.400 | 280 (8) |
| Surinam | 54.760 | 25.320 (46) |
| 60-69 | 35.380 | 14.700 (42) |
| 70-79 | 14.840 | 8.150 (55) |
| 80+ | 4.540 | 2.470 (54) |

| | | |
|---|---|---|
| The Dutch Caribbean | 16.900 | 8.540 (51) |
| 60-69 | 10.620 | 4.860 (46) |
| 70-79 | 5.160 | 3.040 (59) |
| 80+ | 1.130 | 640 (57) |
| Indonesia | 132.190 | 90.650 (69) |
| 60-69 | 72.310 | 45.720 (63) |
| 70-79 | 40.730 | 30.930 (76) |
| 80+ | 19.150 | 14.000 (73) |
| Other Africa | 19.450 | 6.920 (36) |
| 60-69 | 14.990 | 4.920 (33) |
| 70-79 | 3.570 | 1.560 (44) |
| 80+ | 880 | 440 (50) |
| Other Asia | 46.110 | 17.690 (38) |
| 60-69 | 32.260 | 11.290 (35) |
| 70-79 | 11.010 | 5.080 (46) |
| 80+ | 2.850 | 1.320 (46) |
| Other America/Oceania | 20.500 | 12.090 (59) |
| 60-69 | 14.950 | 8.350 (56) |
| 70-79 | 4.510 | 3.070 (68) |
| 80+ | 1.040 | 680 (66) |
| Middle and Eastern European countries within the EU | 18.640 | 9.560 (51) |
| 60-69 | 10.850 | 4.510 (42) |
| 70-79 | 6.100 | 3.970 (65) |
| 80+ | 1.690 | 1.070 (64) |
| GIPS-countries | 13.720 | 7.270 (53) |
| 60-69 | 6.520 | 2.960 (45) |
| 70-79 | 4.690 | 2.780 (59) |
| 80+ | 2.510 | 1.540 (61) |
| Former or associated member states of the Commonwealth of Independent States | 4.870 | 2.410 (50) |
| 60-69 | 2.810 | 1.160 (41) |
| 70-79 | 1.720 | 1.090 (63) |
| 80+ | 340 | 170 (49) |
| Other countries of the EU | 182.150 | 125.750 (69) |
| 60-69 | 62.200 | 37.780 (61) |
| 70-79 | 76.840 | 57.130 (74) |
| 80+ | 43.110 | 30.830 (72) |
| Other European countries | 25.670 | 14.590 (57) |
| 60-69 | 14.590 | 7.400 (51) |
| 70-79 | 8.990 | 5.810 (65) |
| 80+ | 2.100 | 1.380 (66) |
| **Migration status** | | |

| | | |
|---|---|---|
| Born in The Netherlands with both parents born in The Netherlands | 3.434.940 | 2.384.400 (69) |
| 60-69 | 1.615.470 | 1.011.060 (63) |
| 70-79 | 1.253.190 | 959.850 (77) |
| 80+ | 566.280 | 413.500 (73) |
| Born in The Netherlands with one parent born abroad | 188.020 | 132.190 (70) |
| 60-69 | 79.710 | 50.940 (64) |
| 70-79 | 72.630 | 55.280 (76) |
| 80+ | 35.680 | 25.980 (73) |
| Born in The Netherlands with two parents born abroad | 31.560 | 20.160 (64) |
| 60-69 | 22.400 | 13.380 (60) |
| 70-79 | 6.090 | 4.630 (76) |
| 80+ | 3.070 | 2.160 (70) |
| Born abroad with one parent born abroad | 24.530 | 17.200 (70) |
| 60-69 | 8.930 | 5.620 (63) |
| 70-79 | 10.590 | 7.930 (75) |
| 80+ | 5.010 | 3.650 (73) |
| Born abroad with two parents born abroad | 317.350 | 138.170 (44) |
| 60-69 | 181.670 | 66.950 (37) |
| 70-79 | 98.780 | 51.420 (52) |
| 80+ | 36.890 | 19.800 (54) |
| Born abroad with two parents born in The Netherlands | 27.230 | 19.230 (71) |
| 60-69 | 15.010 | 9.710 (65) |
| 70-79 | 7.210 | 5.720 (79) |
| 80+ | 5.010 | 3.810 (76) |
| **Household type** | | |
| One-person household | 1.139.970 | 731.420 (64) |
| 60-69 | 415.010 | 225.530 (54) |
| 70-79 | 419.580 | 290.720 (69) |
| 80+ | 305.370 | 215.160 (70) |
| Unmarried couple without children | 188.780 | 123.720 (66) |
| 60-69 | 122.580 | 73.760 (60) |
| 70-79 | 53.600 | 40.430 (75) |
| 80+ | 12.600 | 9.530 (76) |
| Married couple without children | 2.212.680 | 1.594.170 (72) |
| 60-69 | 1.108.650 | 723.310 (65) |
| 70-79 | 874.510 | 691.400 (79) |
| 80+ | 229.530 | 179.460 (78) |
| Unmarried couple with children | 28.100 | 13.240 (47) |
| 60-69 | 22.730 | 10.170 (45) |
| 70-79 | 4.490 | 2.560 (57) |
| 80+ | 880 | 510 (57) |
| Married couple with children | 237.770 | 124.150 (52) |

| | | |
|---|---|---|
| 60-69 | 183.170 | 89.810 (49) |
| 70-79 | 43.840 | 27.610 (63) |
| 80+ | 10.770 | 6.730 (62) |
| One-parent family | 72.560 | 34.680 (48) |
| 60-69 | 42.290 | 17.460 (41) |
| 70-79 | 18.110 | 9.980 (55) |
| 80+ | 12.160 | 7.240 (60) |
| Other household | 14.530 | 9.280 (64) |
| 60-69 | 8.640 | 5.000 (58) |
| 70-79 | 4.410 | 3.220 (73) |
| 80+ | 1.480 | 1.060 (72) |
| Institutional household | 129.240 | 80.700 (62) |
| 60-69 | 20.140 | 12.600 (63) |
| 70-79 | 29.950 | 18.890 (63) |
| 80+ | 79.150 | 49.200 (62) |
| **Medical risk groups** | | |
| Low medical risk | 2.277.270 | 1.518.980 (67) |
| 18-29 | . | . |
| 30-39 | . | . |
| 40-49 | . | . |
| 50-59 | . | . |
| 60-69 | 1.232.110 | 735.820 (60) |
| 70-79 | 767.150 | 578.860 (75) |
| 80+ | 278.010 | 204.310 (73) |
| Intermediate medical risk | 1.585.720 | 1.080.990 (68) |
| 18-29 | . | . |
| 30-39 | . | . |
| 40-49 | . | . |
| 50-59 | . | . |
| 60-69 | 617.810 | 375.250 (61) |
| 70-79 | 617.270 | 457.930 (74) |
| 80+ | 350.640 | 247.820 (71) |
| High medical risk | 160.640 | 111.380 (69) |
| 18-29 | . | . |
| 30-39 | . | . |
| 40-49 | . | . |
| 50-59 | . | . |
| 60-69 | 73.270 | 46.580 (64) |
| 70-79 | 64.080 | 48.030 (75) |
| 80+ | 23.290 | 16.770 (72) |
| **Socioeconomic position** | | |
| In employment | 628.760 | 356.630 (57) |
| 60-69 | 616.160 | 347.700 (56) |
| 70-79 | 11.400 | 8.080 (71) |

| | | |
|---|---|---|
| 80+ | 1.200 | 850 (71) |
| Self-employed | 149.100 | 79.190 (53) |
| 60-69 | 129.000 | 65.830 (51) |
| 70-79 | 18.230 | 12.060 (66) |
| 80+ | 1.870 | 1.290 (69) |
| Unemployment benefits (WW) | 21.570 | 12.740 (59) |
| 60-69 | 21.550 | 12.720 (59) |
| 70-79 | 20 | . |
| 80+ | . | . |
| Social assistance benefit | 45.040 | 12.530 (28) |
| 60-69 | 43.180 | 12.140 (28) |
| 70-79 | 1.240 | 230 (19) |
| 80+ | 620 | 160 (26) |
| Other benefits | 32.230 | 16.780 (52) |
| 60-69 | 31.430 | 16.230 (52) |
| 70-79 | 270 | 180 (66) |
| 80+ | 530 | 370 (70) |
| Disability benefit | 130.820 | 68.100 (52) |
| 60-69 | 130.800 | 68.090 (52) |
| 70-79 | 20 | . |
| 80+ | . | . |
| Pensioner | 2.911.310 | 2.106.710 (72) |
| 60-69 | 850.050 | 578.490 (68) |
| 70-79 | 1.414.470 | 1.062.530 (75) |
| 80+ | 646.790 | 465.690 (72) |
| Student | 140 | 70 (51) |
| 60-69 | 130 | 60 (49) |
| 70-79 | . | . |
| 80+ | . | . |
| Other/unknown | 104.680 | 58.610 (56) |
| 60-69 | 100.890 | 56.380 (56) |
| 70-79 | 2.850 | 1.700 (60) |
| 80+ | 940 | 530 (56) |
| **Personal income** | | |
| Unknown | 185.210 | 110.140 (59) |
| 60-69 | 117.750 | 67.140 (57) |
| 70-79 | 21.060 | 13.560 (64) |
| 80+ | 46.400 | 29.450 (63) |
| 0 till 10 | 64.320 | 35.770 (56) |
| 60-69 | 61.070 | 33.830 (55) |
| 70-79 | 2.540 | 1.510 (59) |
| 80+ | 710 | 430 (60) |
| 10 till 25 | 820.900 | 559.680 (68) |
| 60-69 | 274.510 | 160.620 (59) |

| | | |
|---|---|---|
| 70-79 | 410.540 | 301.150 (73) |
| 80+ | 135.850 | 97.910 (72) |
| 25 till 50 | 1.313.090 | 861.190 (66) |
| 60-69 | 470.810 | 266.110 (57) |
| 70-79 | 543.810 | 388.420 (71) |
| 80+ | 298.460 | 206.660 (69) |
| 50 till 75 | 936.080 | 652.050 (70) |
| 60-69 | 489.560 | 299.480 (61) |
| 70-79 | 321.520 | 255.570 (79) |
| 80+ | 125.010 | 97.000 (78) |
| 75 till 90 | 424.840 | 294.920 (69) |
| 60-69 | 290.960 | 183.840 (63) |
| 70-79 | 101.340 | 84.480 (83) |
| 80+ | 32.540 | 26.600 (82) |
| 90 till 100 | 279.200 | 197.610 (71) |
| 60-69 | 218.550 | 146.630 (67) |
| 70-79 | 47.680 | 40.140 (84) |
| 80+ | 12.980 | 10.850 (84) |
| **Urbanization** | | |
| Unknown | 1.000 | 630 (63) |
| 60-69 | 400 | 200 (51) |
| 70-79 | 270 | 190 (71) |
| 80+ | 330 | 230 (70) |
| Not urbanized | 706.280 | 464.940 (66) |
| 60-69 | 361.870 | 212.200 (59) |
| 70-79 | 248.610 | 184.750 (74) |
| 80+ | 95.800 | 67.990 (71) |
| Hardly urbanized | 700.420 | 486.000 (69) |
| 60-69 | 328.990 | 204.910 (62) |
| 70-79 | 257.170 | 197.530 (77) |
| 80+ | 114.270 | 83.560 (73) |
| Moderately urbanized | 794.100 | 553.380 (70) |
| 60-69 | 375.540 | 235.010 (63) |
| 70-79 | 290.610 | 224.100 (77) |
| 80+ | 127.950 | 94.270 (74) |
| Strongly urbanized | 1.054.420 | 717.400 (68) |
| 60-69 | 488.510 | 297.070 (61) |
| 70-79 | 382.690 | 287.580 (75) |
| 80+ | 183.220 | 132.750 (72) |
| Extremely urbanized | 767.410 | 489.010 (64) |
| 60-69 | 367.890 | 208.250 (57) |
| 70-79 | 269.140 | 190.660 (71) |
| 80+ | 130.370 | 90.100 (69) |
| **Long term care recipients, residential, nursing home** | | |

| | | |
|---|---|---|
| No | 3.916.440 | 2.646.440 (68) |
| Yes | 107.190 | 64.920 (61) |
| **Long term care recipients, residential, mentally impaired** | | |
| 0 | 4.008.590 | 2.700.840 (67) |
| 1 | 15.040 | 10.520 (70) |
| **Long term care recipients, non-residential, mentally impaired** | | |
| 0 | 4.022.020 | 2.710.470 (67) |
| 1 | 1.610 | 880 (55) |
| **Voting proportions political movements (%)** | | |
| **Progressive liberal** | | |
| 0-0.1 | 419.130 | 257.720 (61) |
| 60-69 | 202.340 | 109.050 (54) |
| 70-79 | 151.080 | 105.090 (70) |
| 80+ | 65.720 | 43.590 (66) |
| 0.1-0.2 | 2.590.160 | 1.752.870 (68) |
| 60-69 | 1.232.940 | 743.050 (60) |
| 70-79 | 939.160 | 707.630 (75) |
| 80+ | 418.060 | 302.190 (72) |
| 0.2-0.3 | 686.670 | 476.550 (69) |
| 60-69 | 319.770 | 200.180 (63) |
| 70-79 | 244.900 | 186.800 (76) |
| 80+ | 122.000 | 89.580 (73) |
| 0.3-0.4 | 189.490 | 128.880 (68) |
| 60-69 | 96.810 | 60.900 (63) |
| 70-79 | 65.680 | 48.770 (74) |
| 80+ | 27.000 | 19.210 (71) |
| 0.4-0.5 | 21.170 | 15.830 (75) |
| 60-69 | 11.420 | 8.100 (71) |
| 70-79 | 7.150 | 5.770 (81) |
| 80+ | 2.600 | 1.960 (75) |
| Unknown | 117.000 | 79.500 (68) |
| 60-69 | 59.930 | 36.360 (61) |
| 70-79 | 40.510 | 30.770 (76) |
| 80+ | 16.570 | 12.370 (75) |
| **Right-wing liberal** | | |
| 0-0.1 | 55.920 | 28.050 (50) |
| 60-69 | 31.990 | 14.350 (45) |
| 70-79 | 17.690 | 10.100 (57) |
| 80+ | 6.230 | 3.600 (58) |
| 0.1-0.2 | 1.360.630 | 867.470 (64) |
| 60-69 | 655.320 | 370.600 (57) |
| 70-79 | 486.090 | 346.260 (71) |
| 80+ | 219.220 | 150.620 (69) |
| 0.2-0.3 | 2.107.960 | 1.465.170 (70) |

| | | |
|---|---|---|
| 60-69 | 996.480 | 621.980 (62) |
| 70-79 | 765.530 | 588.660 (77) |
| 80+ | 345.950 | 254.530 (74) |
| 0.3-0.4 | 379.860 | 269.490 (71) |
| 60-69 | 178.640 | 113.800 (64) |
| 70-79 | 137.820 | 108.350 (79) |
| 80+ | 63.400 | 47.340 (75) |
| 0.4-0.5 | 2.260 | 1.690 (75) |
| 60-69 | 850 | 570 (67) |
| 70-79 | 850 | 680 (80) |
| 80+ | 560 | 440 (79) |
| Unknown | 117.000 | 79.500 (68) |
| 60-69 | 59.930 | 36.360 (61) |
| 70-79 | 40.510 | 30.770 (76) |
| 80+ | 16.570 | 12.370 (75) |
| **Right-wing Christian** | | |
| 0-0.1 | 3.728.080 | 2.517.310 (68) |
| 60-69 | 1.779.190 | 1.073.240 (60) |
| 70-79 | 1.342.610 | 1.007.270 (75) |
| 80+ | 606.280 | 436.800 (72) |
| 0.1-0.2 | 104.820 | 69.900 (67) |
| 60-69 | 48.330 | 28.870 (60) |
| 70-79 | 38.680 | 28.620 (74) |
| 80+ | 17.800 | 12.410 (70) |
| 0.2-0.3 | 44.760 | 28.070 (63) |
| 60-69 | 20.810 | 11.510 (55) |
| 70-79 | 16.560 | 11.670 (70) |
| 80+ | 7.380 | 4.890 (66) |
| 0.3-0.4 | 9.860 | 5.020 (51) |
| 60-69 | 5.020 | 2.260 (45) |
| 70-79 | 3.490 | 2.030 (58) |
| 80+ | 1.350 | 720 (54) |
| 0.4-0.5 | 8.120 | 4.740 (58) |
| 60-69 | 4.210 | 2.220 (53) |
| 70-79 | 2.780 | 1.790 (64) |
| 80+ | 1.130 | 730 (64) |
| 0.5-0.6 | 3.000 | 1.250 (42) |
| 60-69 | 1.550 | 570 (37) |
| 70-79 | 1.070 | 500 (46) |
| 80+ | 380 | 180 (47) |
| 0.7+ | 40 | 20 (60) |
| 60-69 | 10 | . |
| 70-79 | 10 | . |
| 80+ | . | . |

| | | |
|---|---|---|
| Unknown | 124.970 | 85.060 (68) |
| 60-69 | 64.080 | 38.970 (61) |
| 70-79 | 43.280 | 32.930 (76) |
| 80+ | 17.620 | 13.160 (75) |
| **Christian middle** | | |
| 0-0.1 | 1.303.450 | 852.010 (65) |
| 60-69 | 639.090 | 373.280 (58) |
| 70-79 | 457.910 | 333.080 (73) |
| 80+ | 206.450 | 145.650 (71) |
| 0.1-0.2 | 2.099.450 | 1.451.820 (69) |
| 60-69 | 983.180 | 608.950 (62) |
| 70-79 | 768.400 | 588.260 (77) |
| 80+ | 347.870 | 254.600 (73) |
| 0.2-0.3 | 409.020 | 268.220 (66) |
| 60-69 | 195.340 | 113.900 (58) |
| 70-79 | 147.640 | 108.360 (73) |
| 80+ | 66.040 | 45.970 (70) |
| 0.3-0.4 | 84.760 | 53.860 (64) |
| 60-69 | 40.680 | 22.630 (56) |
| 70-79 | 30.460 | 21.890 (72) |
| 80+ | 13.630 | 9.340 (69) |
| 0.4-0.5 | 9.960 | 5.950 (60) |
| 60-69 | 4.990 | 2.530 (51) |
| 70-79 | 3.590 | 2.470 (69) |
| 80+ | 1.390 | 960 (69) |
| Unknown | 117.000 | 79.500 (68) |
| 60-69 | 59.930 | 36.360 (61) |
| 70-79 | 40.510 | 30.770 (76) |
| 80+ | 16.570 | 12.370 (75) |
| **Right-wing conservative** | | |
| 0-0.1 | 206.670 | 138.730 (67) |
| 60-69 | 105.400 | 64.590 (61) |
| 70-79 | 70.760 | 52.250 (74) |
| 80+ | 30.510 | 21.890 (72) |
| 0.1-0.2 | 1.638.340 | 1.132.150 (69) |
| 60-69 | 767.960 | 477.520 (62) |
| 70-79 | 591.990 | 451.330 (76) |
| 80+ | 278.390 | 203.300 (73) |
| 0.2-0.3 | 1.797.470 | 1.199.710 (67) |
| 60-69 | 859.510 | 509.670 (59) |
| 70-79 | 651.330 | 485.690 (75) |
| 80+ | 286.640 | 204.360 (71) |
| 0.3-0.4 | 248.550 | 153.510 (62) |
| 60-69 | 122.100 | 66.000 (54) |

| | | |
|---|---|---|
| 70-79 | 88.470 | 61.610 (70) |
| 80+ | 37.970 | 25.900 (68) |
| 0.4-0.5 | 8.360 | 4.500 (54) |
| 60-69 | 4.640 | 2.130 (46) |
| 70-79 | 2.740 | 1.760 (64) |
| 80+ | 980 | 600 (61) |
| 0.5-0.6 | 7.250 | 3.260 (45) |
| 60-69 | 3.660 | 1.380 (38) |
| 70-79 | 2.700 | 1.410 (52) |
| 80+ | 890 | 470 (53) |
| Unknown | 117.000 | 79.500 (68) |
| 60-69 | 59.930 | 36.360 (61) |
| 70-79 | 40.510 | 30.770 (76) |
| 80+ | 16.570 | 12.370 (75) |
| **Progressive left-wing** | | |
| 0-0.1 | 120.420 | 72.060 (60) |
| 60-69 | 58.270 | 30.580 (52) |
| 70-79 | 43.110 | 29.110 (68) |
| 80+ | 19.030 | 12.380 (65) |
| 0.1-0.2 | 1.560.610 | 1.075.600 (69) |
| 60-69 | 743.070 | 458.270 (62) |
| 70-79 | 566.630 | 434.060 (77) |
| 80+ | 250.910 | 183.270 (73) |
| 0.2-0.3 | 1.840.780 | 1.237.200 (67) |
| 60-69 | 865.100 | 518.380 (60) |
| 70-79 | 665.730 | 496.400 (75) |
| 80+ | 309.960 | 222.420 (72) |
| 0.3-0.4 | 339.980 | 221.040 (65) |
| 60-69 | 169.990 | 99.860 (59) |
| 70-79 | 118.710 | 85.450 (72) |
| 80+ | 51.290 | 35.730 (70) |
| 0.4-0.5 | 44.840 | 25.960 (58) |
| 60-69 | 26.850 | 14.200 (53) |
| 70-79 | 13.800 | 9.040 (65) |
| 80+ | 4.190 | 2.720 (65) |
| Unknown | 117.000 | 79.500 (68) |
| 60-69 | 59.930 | 36.360 (61) |
| 70-79 | 40.510 | 30.770 (76) |
| 80+ | 16.570 | 12.370 (75) |

**Supplemental Table 3. Autumn booster uptake per determinant and age group for individuals with a medical risk, aged <60 years.**

| | N | Autumn booster uptake (N, %) |
|---|---|---|
| **Age** | | |
| 18-29 | 217.790 | 32.830 (15) |
| 30-39 | 220.580 | 45.940 (21) |
| 40-49 | 328.080 | 92.210 (28) |
| 50-59 | 573.950 | 230.300 (40) |
| **Sex** | | |
| Male | 616.070 | 182.070 (30) |
| 18-29 | 101.280 | 14.280 (14) |
| 30-39 | 94.650 | 19.350 (20) |
| 40-49 | 143.790 | 39.560 (28) |
| 50-59 | 275.340 | 108.880 (39) |
| Female | 724.340 | 219.210 (30) |
| 18-29 | 116.510 | 18.540 (16) |
| 30-39 | 125.930 | 26.590 (21) |
| 40-49 | 184.280 | 52.660 (29) |
| 50-59 | 297.610 | 121.420 (41) |
| **Household car ownership** | | |
| Yes | 1.053.530 | 310.480 (29) |
| 18-29 | 146.730 | 17.760 (12) |
| 30-39 | 166.780 | 30.260 (18) |
| 40-49 | 264.240 | 70.960 (27) |
| 50-59 | 475.780 | 191.500 (40) |
| No | 286.870 | 90.790 (32) |
| 18-29 | 71.070 | 15.070 (21) |
| 30-39 | 53.800 | 15.670 (29) |
| 40-49 | 63.840 | 21.250 (33) |
| 50-59 | 98.170 | 38.800 (40) |
| **Employment sector** | | |
| Activities of household as employer, undifferentiated goods and service producing activities of household for own use | 3.860 | 860 (22) |
| 18-29 | 240 | 20 (9) |
| 30-39 | 450 | 50 (11) |
| 40-49 | 1.150 | 190 (17) |
| 50-59 | 2.020 | 600 (30) |
| Agriculture, forestry and fishery | 6.580 | 1.430 (22) |
| 18-29 | 1.320 | 90 (7) |
| 30-39 | 920 | 100 (11) |
| 40-49 | 1.560 | 320 (20) |
| 50-59 | 2.780 | 920 (33) |
| Mining and quarrying | 800 | 260 (33) |

| | | |
|---|---|---|
| 18-29 | 70 | . |
| 30-39 | 120 | 20 (19) |
| 40-49 | 250 | 70 (29) |
| 50-59 | 360 | 160 (43) |
| Industry | 93.460 | 28.370 (30) |
| 18-29 | 9.580 | 1.130 (12) |
| 30-39 | 13.790 | 2.710 (20) |
| 40-49 | 23.710 | 6.510 (27) |
| 50-59 | 46.370 | 18.020 (39) |
| Electricity supply | 3.610 | 1.200 (33) |
| 18-29 | 390 | 50 (13) |
| 30-39 | 740 | 160 (21) |
| 40-49 | 1.020 | 330 (32) |
| 50-59 | 1.460 | 670 (46) |
| Water supply, sewage and waste management | 4.670 | 1.260 (27) |
| 18-29 | 420 | 40 (9) |
| 30-39 | 650 | 80 (12) |
| 40-49 | 1.200 | 270 (22) |
| 50-59 | 2.400 | 880 (37) |
| Construction | 37.140 | 8.310 (22) |
| 18-29 | 5.090 | 290 (6) |
| 30-39 | 5.920 | 600 (10) |
| 40-49 | 10.040 | 2.040 (20) |
| 50-59 | 16.090 | 5.390 (33) |
| Wholesail and retail trade | 119.810 | 28.300 (24) |
| 18-29 | 25.710 | 2.270 (9) |
| 30-39 | 20.090 | 2.890 (14) |
| 40-49 | 29.880 | 6.980 (23) |
| 50-59 | 44.130 | 16.150 (37) |
| Transportation and storage | 43.170 | 11.570 (27) |
| 18-29 | 5.020 | 380 (8) |
| 30-39 | 5.990 | 830 (14) |
| 40-49 | 10.650 | 2.450 (23) |
| 50-59 | 21.520 | 7.920 (37) |
| Accommodation and food services activities | 25.870 | 4.430 (17) |
| 18-29 | 9.890 | 780 (8) |
| 30-39 | 3.860 | 500 (13) |
| 40-49 | 4.910 | 950 (19) |
| 50-59 | 7.220 | 2.200 (30) |
| Information and communication | 29.230 | 9.790 (33) |
| 18-29 | 6.240 | 980 (16) |
| 30-39 | 7.410 | 1.840 (25) |
| 40-49 | 7.230 | 2.600 (36) |
| 50-59 | 8.350 | 4.370 (52) |

| | | |
|---|---|---|
| Financial services | 29.450 | 10.360 (35) |
| 18-29 | 2.880 | 400 (14) |
| 30-39 | 5.370 | 1.180 (22) |
| 40-49 | 8.770 | 2.830 (32) |
| 50-59 | 12.430 | 5.950 (48) |
| Real estate activities | 7.550 | 2.260 (30) |
| 18-29 | 970 | 100 (10) |
| 30-39 | 1.260 | 190 (15) |
| 40-49 | 2.090 | 580 (28) |
| 50-59 | 3.240 | 1.400 (43) |
| Professional scientific and technical activities | 52.740 | 16.920 (32) |
| 18-29 | 11.100 | 1.680 (15) |
| 30-39 | 10.660 | 2.490 (23) |
| 40-49 | 13.530 | 4.400 (32) |
| 50-59 | 17.450 | 8.360 (48) |
| Administrative and support service activities | 74.430 | 15.720 (21) |
| 18-29 | 17.820 | 1.750 (10) |
| 30-39 | 12.200 | 1.650 (14) |
| 40-49 | 17.220 | 3.530 (21) |
| 50-59 | 27.180 | 8.790 (32) |
| Public administration and defence | 65.830 | 24.760 (38) |
| 18-29 | 6.070 | 1.010 (17) |
| 30-39 | 10.760 | 2.410 (22) |
| 40-49 | 17.900 | 5.970 (33) |
| 50-59 | 31.100 | 15.380 (49) |
| Education | 63.440 | 21.950 (35) |
| 18-29 | 9.560 | 1.640 (17) |
| 30-39 | 13.690 | 3.230 (24) |
| 40-49 | 17.320 | 5.700 (33) |
| 50-59 | 22.860 | 11.380 (50) |
| Human health and social work activities | 187.770 | 61.050 (33) |
| 18-29 | 33.450 | 5.030 (15) |
| 30-39 | 36.060 | 8.150 (23) |
| 40-49 | 45.710 | 14.960 (33) |
| 50-59 | 72.560 | 32.910 (45) |
| Arts, entertainment and recreation | 11.900 | 3.130 (26) |
| 18-29 | 3.390 | 360 (11) |
| 30-39 | 1.930 | 390 (20) |
| 40-49 | 2.530 | 670 (27) |
| 50-59 | 4.050 | 1.710 (42) |
| Other service activities | 12.540 | 3.810 (30) |
| 18-29 | 2.330 | 300 (13) |
| 30-39 | 2.200 | 410 (18) |
| 40-49 | 3.170 | 920 (29) |

| | | |
|---|---|---|
| 50-59 | 4.850 | 2.180 (45) |
| Activities of extraterritorial organisations and bodies | 120 | 30 (22) |
| 18-29 | . | . |
| 30-39 | 20 | . |
| 40-49 | 30 | . |
| 50-59 | 60 | 20 (26) |
| Other/unemployed/unknown | 466.450 | 145.500 (31) |
| 18-29 | 66.250 | 14.530 (22) |
| 30-39 | 66.510 | 16.060 (24) |
| 40-49 | 108.210 | 29.940 (28) |
| 50-59 | 225.480 | 84.960 (38) |
| **Country of origin** | | |
| The Netherlands | 1.020.850 | 333.000 (33) |
| 18-29 | 172.230 | 27.160 (16) |
| 30-39 | 159.820 | 36.220 (23) |
| 40-49 | 240.790 | 75.040 (31) |
| 50-59 | 448.010 | 194.580 (43) |
| Turkey | 38.380 | 2.620 (7) |
| 18-29 | 4.570 | 240 (5) |
| 30-39 | 6.140 | 340 (6) |
| 40-49 | 11.620 | 730 (6) |
| 50-59 | 16.060 | 1.310 (8) |
| Morocco | 25.890 | 1.350 (5) |
| 18-29 | 3.310 | 140 (4) |
| 30-39 | 4.450 | 210 (5) |
| 40-49 | 7.990 | 430 (5) |
| 50-59 | 10.140 | 560 (6) |
| Surinam | 38.330 | 8.680 (23) |
| 18-29 | 4.710 | 550 (12) |
| 30-39 | 6.420 | 1.000 (16) |
| 40-49 | 9.970 | 2.100 (21) |
| 50-59 | 17.230 | 5.030 (29) |
| The Dutch Caribbean | 12.060 | 2.820 (23) |
| 18-29 | 2.290 | 300 (13) |
| 30-39 | 2.430 | 420 (17) |
| 40-49 | 3.130 | 740 (24) |
| 50-59 | 4.210 | 1.360 (32) |
| Indonesia | 33.450 | 12.750 (38) |
| 18-29 | 1.600 | 330 (21) |
| 30-39 | 4.150 | 1.000 (24) |
| 40-49 | 8.950 | 2.800 (31) |
| 50-59 | 18.750 | 8.620 (46) |
| Other Africa | 22.600 | 3.750 (17) |
| 18-29 | 3.910 | 430 (11) |

| | | |
|---|---|---|
| 30-39 | 4.880 | 630 (13) |
| 40-49 | 6.150 | 1.000 (16) |
| 50-59 | 7.670 | 1.690 (22) |
| Other Asia | 47.440 | 8.550 (18) |
| 18-29 | 7.760 | 960 (12) |
| 30-39 | 11.400 | 1.750 (15) |
| 40-49 | 12.980 | 2.400 (18) |
| 50-59 | 15.310 | 3.440 (23) |
| Other America/Oceania | 19.540 | 5.750 (29) |
| 18-29 | 4.310 | 790 (18) |
| 30-39 | 5.070 | 1.350 (27) |
| 40-49 | 4.810 | 1.460 (30) |
| 50-59 | 5.350 | 2.150 (40) |
| Middle and eastern European countries within the EU | 15.390 | 2.260 (15) |
| 18-29 | 2.510 | 260 (10) |
| 30-39 | 4.200 | 460 (11) |
| 40-49 | 4.700 | 640 (14) |
| 50-59 | 3.970 | 900 (23) |
| GIPS countries | 11.610 | 3.040 (26) |
| 18-29 | 1.840 | 230 (13) |
| 30-39 | 2.350 | 520 (22) |
| 40-49 | 3.250 | 840 (26) |
| 50-59 | 4.160 | 1.440 (35) |
| Former or associated member states of the Commonwealth of Independent States | 2.570 | 450 (17) |
| 18-29 | 460 | 40 (9) |
| 30-39 | 730 | 130 (17) |
| 40-49 | 730 | 130 (17) |
| 50-59 | 650 | 160 (24) |
| Other countries of the EU | 37.910 | 12.520 (33) |
| 18-29 | 5.740 | 970 (17) |
| 30-39 | 5.740 | 1.360 (24) |
| 40-49 | 9.210 | 2.900 (31) |
| 50-59 | 17.230 | 7.300 (42) |
| Other European countries | 14.400 | 3.760 (26) |
| 18-29 | 2.560 | 420 (16) |
| 30-39 | 2.810 | 560 (20) |
| 40-49 | 3.810 | 1.010 (27) |
| 50-59 | 5.210 | 1.770 (34) |
| **Migration status** | | |
| Born in The Netherlands with both parents born in The Netherlands | 1.020.850 | 333.000 (33) |
| 18-29 | 172.230 | 27.160 (16) |

| | | |
|---|---|---|
| 30-39 | 159.820 | 36.220 (23) |
| 40-49 | 240.790 | 75.040 (31) |
| 50-59 | 448.010 | 194.580 (43) |
| Born in the Netherlands with one parent born abroad | 71.050 | 21.930 (31) |
| 18-29 | 15.620 | 2.460 (16) |
| 30-39 | 12.540 | 2.760 (22) |
| 40-49 | 16.130 | 4.980 (31) |
| 50-59 | 26.750 | 11.720 (44) |
| Born in the Netherlands with two parents born abroad | 44.170 | 7.870 (18) |
| 18-29 | 13.200 | 1.090 (8) |
| 30-39 | 12.630 | 1.470 (12) |
| 40-49 | 10.420 | 1.980 (19) |
| 50-59 | 7.930 | 3.320 (42) |
| Born abroad with one parent born abroad | 7.490 | 2.390 (32) |
| 18-29 | 1.320 | 240 (18) |
| 30-39 | 1.320 | 350 (27) |
| 40-49 | 1.980 | 600 (30) |
| 50-59 | 2.880 | 1.200 (41) |
| Born abroad with two parents born abroad | 184.080 | 31.690 (17) |
| 18-29 | 12.810 | 1.290 (10) |
| 30-39 | 31.600 | 4.350 (14) |
| 40-49 | 55.200 | 8.380 (15) |
| 50-59 | 84.470 | 17.670 (21) |
| Born abroad with two parents born in the Netherlands | 12.770 | 4.420 (35) |
| 18-29 | 2.620 | 580 (22) |
| 30-39 | 2.670 | 790 (29) |
| 40-49 | 3.560 | 1.240 (35) |
| 50-59 | 3.910 | 1.820 (46) |
| **Household type** | | |
| One-person household | 243.550 | 74.230 (30) |
| 18-29 | 48.040 | 7.380 (15) |
| 30-39 | 36.580 | 8.580 (23) |
| 40-49 | 49.440 | 15.610 (32) |
| 50-59 | 109.490 | 42.660 (39) |
| Unmarried couple without children | 116.640 | 32.810 (28) |
| 18-29 | 39.110 | 5.640 (14) |
| 30-39 | 19.660 | 4.490 (23) |
| 40-49 | 17.660 | 5.820 (33) |
| 50-59 | 40.210 | 16.870 (42) |
| Married couple without children | 208.500 | 91.560 (44) |
| 18-29 | 8.960 | 1.340 (15) |
| 30-39 | 12.590 | 3.550 (28) |

| | | |
|---|---|---|
| 40-49 | 25.090 | 9.760 (39) |
| 50-59 | 161.860 | 76.920 (48) |
| Unmarried couple with children | 113.380 | 22.970 (20) |
| 18-29 | 13.970 | 1.020 (7) |
| 30-39 | 32.190 | 4.350 (14) |
| 40-49 | 38.640 | 8.540 (22) |
| 50-59 | 28.590 | 9.050 (32) |
| Married couple with children | 482.220 | 122.980 (26) |
| 18-29 | 68.630 | 8.050 (12) |
| 30-39 | 87.970 | 15.210 (17) |
| 40-49 | 149.170 | 37.740 (25) |
| 50-59 | 176.440 | 61.990 (35) |
| One-parent family | 109.200 | 21.140 (19) |
| 18-29 | 20.040 | 1.740 (9) |
| 30-39 | 17.280 | 1.970 (11) |
| 40-49 | 34.370 | 6.670 (19) |
| 50-59 | 37.510 | 10.770 (29) |
| Other household | 7.580 | 1.810 (24) |
| 18-29 | 3.080 | 380 (12) |
| 30-39 | 1.040 | 160 (15) |
| 40-49 | 990 | 260 (26) |
| 50-59 | 2.460 | 1.020 (41) |
| Institutional household | 59.340 | 33.760 (57) |
| 18-29 | 15.960 | 7.270 (46) |
| 30-39 | 13.280 | 7.630 (57) |
| 40-49 | 12.710 | 7.830 (62) |
| 50-59 | 17.390 | 11.030 (63) |
| **Medical risk groups** | | |
| Low medical risk | 67.890 | 33.490 (49) |
| 18-29 | 23.600 | 8.420 (36) |
| 30-39 | 15.770 | 7.860 (50) |
| 40-49 | 13.070 | 7.610 (58) |
| 50-59 | 15.450 | 9.590 (62) |
| Intermediate medical risk | 1.130.540 | 320.050 (28) |
| 18-29 | 179.250 | 22.020 (12) |
| 30-39 | 182.110 | 33.160 (18) |
| 40-49 | 276.370 | 72.660 (26) |
| 50-59 | 492.810 | 192.220 (39) |
| High medical risk | 141.970 | 47.740 (34) |
| 18-29 | 14.950 | 2.380 (16) |
| 30-39 | 22.710 | 4.920 (22) |
| 40-49 | 38.640 | 11.950 (31) |
| 50-59 | 65.680 | 28.490 (43) |
| **Socioeconomic position** | | |

| | | |
|---|---|---|
| In employment | 823.860 | 248.640 (30) |
| 18-29 | 111.200 | 13.410 (12) |
| 30-39 | 152.140 | 29.420 (19) |
| 40-49 | 215.980 | 61.400 (28) |
| 50-59 | 344.550 | 144.410 (42) |
| Self-employed | 105.150 | 26.400 (25) |
| 18-29 | 6.390 | 490 (8) |
| 30-39 | 16.460 | 2.170 (13) |
| 40-49 | 30.180 | 6.170 (20) |
| 50-59 | 52.120 | 17.580 (34) |
| Unemployment benefits (WW) | 10.930 | 2.990 (27) |
| 18-29 | 520 | 60 (11) |
| 30-39 | 1.630 | 180 (11) |
| 40-49 | 3.050 | 660 (22) |
| 50-59 | 5.730 | 2.090 (36) |
| Social assistance benefit | 65.240 | 11.830 (18) |
| 18-29 | 5.640 | 750 (13) |
| 30-39 | 8.760 | 870 (10) |
| 40-49 | 17.390 | 2.640 (15) |
| 50-59 | 33.450 | 7.570 (23) |
| Other benefits | 88.930 | 40.360 (45) |
| 18-29 | 26.460 | 9.720 (37) |
| 30-39 | 23.490 | 9.890 (42) |
| 40-49 | 17.820 | 9.240 (52) |
| 50-59 | 21.150 | 11.510 (54) |
| Disability benefit | 120.530 | 41.870 (35) |
| 18-29 | 3.040 | 340 (11) |
| 30-39 | 11.190 | 2.270 (20) |
| 40-49 | 30.280 | 8.850 (29) |
| 50-59 | 76.020 | 30.410 (40) |
| Pensioner | 13.920 | 5.610 (40) |
| 18-29 | 150 | 30 (18) |
| 30-39 | 430 | 80 (18) |
| 40-49 | 2.220 | 640 (29) |
| 50-59 | 11.120 | 4.870 (44) |
| Student | 63.330 | 8.000 (13) |
| 18-29 | 61.690 | 7.620 (12) |
| 30-39 | 920 | 170 (19) |
| 40-49 | 470 | 120 (24) |
| 50-59 | 250 | 90 (35) |
| Unknown | 48.520 | 15.570 (32) |
| 18-29 | 2.720 | 410 (15) |
| 30-39 | 5.550 | 880 (16) |
| 40-49 | 10.700 | 2.500 (23) |

| | | |
|---|---|---|
| 50-59 | 29.560 | 11.790 (40) |
| **Personal income** | | |
| Unknown | 110.400 | 48.980 (44) |
| 18-29 | 21.820 | 7.920 (36) |
| 30-39 | 18.970 | 8.490 (45) |
| 40-49 | 23.410 | 10.210 (44) |
| 50-59 | 46.200 | 22.350 (48) |
| 0 till 10 | 59.250 | 12.220 (21) |
| 18-29 | 30.230 | 3.920 (13) |
| 30-39 | 4.660 | 650 (14) |
| 40-49 | 8.040 | 1.720 (21) |
| 50-59 | 16.330 | 5.930 (36) |
| 10 till 25 | 135.260 | 27.340 (20) |
| 18-29 | 43.240 | 4.730 (11) |
| 30-39 | 14.550 | 1.740 (12) |
| 40-49 | 26.360 | 4.970 (19) |
| 50-59 | 51.110 | 15.910 (31) |
| 25 till 50 | 321.360 | 82.930 (26) |
| 18-29 | 54.690 | 7.210 (13) |
| 30-39 | 55.080 | 9.590 (17) |
| 40-49 | 77.360 | 19.060 (25) |
| 50-59 | 134.230 | 47.070 (35) |
| 50 till 75 | 334.130 | 92.410 (28) |
| 18-29 | 49.160 | 5.790 (12) |
| 30-39 | 62.040 | 10.480 (17) |
| 40-49 | 82.280 | 21.400 (26) |
| 50-59 | 140.660 | 54.750 (39) |
| 75 till 90 | 217.570 | 71.640 (33) |
| 18-29 | 15.460 | 2.760 (18) |
| 30-39 | 41.670 | 9.050 (22) |
| 40-49 | 59.600 | 17.260 (29) |
| 50-59 | 100.840 | 42.570 (42) |
| 90 till 100 | 162.420 | 65.760 (40) |
| 18-29 | 3.190 | 500 (16) |
| 30-39 | 23.610 | 5.940 (25) |
| 40-49 | 51.030 | 17.600 (34) |
| 50-59 | 84.580 | 41.720 (49) |
| **Education level** | | |
| Unknown | 357.710 | 130.300 (36) |
| 18-29 | 6.370 | 1.490 (23) |
| 30-39 | 30.000 | 8.380 (28) |
| 40-49 | 93.970 | 26.620 (28) |
| 50-59 | 227.380 | 93.820 (41) |
| Primary education | 101.610 | 25.420 (25) |

| | | |
|---|---|---|
| 18-29 | 21.850 | 7.560 (35) |
| 30-39 | 12.600 | 3.170 (25) |
| 40-49 | 22.070 | 3.930 (18) |
| 50-59 | 45.100 | 10.760 (24) |
| Prevocational secondary education-basic vocational programme (VMBO-b/k), lower secondary vocational training and assistant's training (MBO-1) | 96.180 | 22.860 (24) |
| 18-29 | 16.950 | 1.790 (11) |
| 30-39 | 14.170 | 2.300 (16) |
| 40-49 | 21.430 | 4.450 (21) |
| 50-59 | 43.630 | 14.320 (33) |
| Prevocational secondary education – theoretical and vocational program (VMBO-g/t), the first three years of senior general secondary education (HAVO) and pre-university secondary education (VWO) | 41.710 | 9.610 (23) |
| 18-29 | 12.070 | 1.260 (10) |
| 30-39 | 6.090 | 1.050 (17) |
| 40-49 | 8.220 | 1.830 (22) |
| 50-59 | 15.330 | 5.480 (36) |
| Basic vocational training (MBO-2) and vocational training (MBO-3) | 163.970 | 35.330 (22) |
| 18-29 | 32.260 | 2.390 (7) |
| 30-39 | 33.730 | 3.990 (12) |
| 40-49 | 36.200 | 7.700 (21) |
| 50-59 | 61.790 | 21.250 (34) |
| Middle management and specialist education (MBO-4) | 166.070 | 40.080 (24) |
| 18-29 | 38.490 | 3.310 (9) |
| 30-39 | 36.060 | 5.400 (15) |
| 40-49 | 38.540 | 9.920 (26) |
| 50-59 | 52.980 | 21.450 (40) |
| Upper secondary education (HAVO/VWO) | 80.130 | 19.460 (24) |
| 18-29 | 35.550 | 4.440 (12) |
| 30-39 | 9.470 | 1.710 (18) |
| 40-49 | 12.870 | 3.580 (28) |
| 50-59 | 22.250 | 9.740 (44) |
| Hbo-, wo-bachelor | 211.820 | 66.860 (32) |
| 18-29 | 38.570 | 6.350 (16) |
| 30-39 | 48.770 | 10.130 (21) |
| 40-49 | 59.190 | 19.360 (33) |
| 50-59 | 65.290 | 31.020 (48) |
| Hbo-, wo-master, doctor | 121.200 | 51.360 (42) |
| 18-29 | 15.700 | 4.260 (27) |
| 30-39 | 29.710 | 9.810 (33) |
| 40-49 | 35.600 | 14.830 (42) |
| 50-59 | 40.200 | 22.470 (56) |
| **Urbanization** | | |
| Unknown | 280 | 60 (21) |

| | | |
|---|---|---|
| 18-29 | 60 | . |
| 30-39 | 50 | . |
| 40-49 | 60 | 10 (17) |
| 50-59 | 110 | 30 (30) |
| Not urbanized | 210.660 | 63.110 (30) |
| 18-29 | 32.050 | 4.650 (15) |
| 30-39 | 30.560 | 6.060 (20) |
| 40-49 | 49.630 | 13.530 (27) |
| 50-59 | 98.420 | 38.870 (39) |
| Hardly urbanized | 216.290 | 68.600 (32) |
| 18-29 | 32.210 | 5.040 (16) |
| 30-39 | 35.590 | 7.550 (21) |
| 40-49 | 53.950 | 15.730 (29) |
| 50-59 | 94.550 | 40.280 (43) |
| Moderately urbanized | 253.060 | 79.860 (32) |
| 18-29 | 36.600 | 5.510 (15) |
| 30-39 | 41.760 | 8.840 (21) |
| 40-49 | 64.080 | 18.890 (29) |
| 50-59 | 110.620 | 46.610 (42) |
| Strongly urbanized | 343.910 | 103.730 (30) |
| 18-29 | 51.370 | 7.610 (15) |
| 30-39 | 56.880 | 11.590 (20) |
| 40-49 | 86.700 | 24.560 (28) |
| 50-59 | 148.970 | 59.960 (40) |
| Extremely urbanized | 316.200 | 85.920 (27) |
| 18-29 | 65.510 | 10.010 (15) |
| 30-39 | 55.750 | 11.890 (21) |
| 40-49 | 73.650 | 19.490 (26) |
| 50-59 | 121.280 | 44.530 (37) |
| **Long term care recipients, residential, nursing home** | | |
| No | 1.338.140 | 400.030 (30) |
| 18-29 | 217.770 | 32.820 (15) |
| 30-39 | 220.490 | 45.890 (21) |
| 40-49 | 327.710 | 92.010 (28) |
| 50-59 | 572.170 | 229.310 (40) |
| Yes | 2.260 | 1.250 (55) |
| 18-29 | 20 | . |
| 30-39 | 100 | 50 (50) |
| 40-49 | 370 | 200 (55) |
| 50-59 | 1.770 | 990 (56) |
| **Long term care recipients, residential, mentally impaired** | | |
| No | 1.287.530 | 370.780 (29) |
| 18-29 | 201.240 | 25.480 (13) |
| 30-39 | 207.870 | 38.540 (19) |

| | | |
|---|---|---|
| 40-49 | 317.400 | 85.230 (27) |
| 50-59 | 561.030 | 221.530 (39) |
| Yes | 52.870 | 30.500 (58) |
| 18-29 | 16.550 | 7.350 (44) |
| 30-39 | 12.720 | 7.390 (58) |
| 40-49 | 10.680 | 6.980 (65) |
| 50-59 | 12.920 | 8.770 (68) |
| **Long term care recipients, non-residential, mentally impaired** | | |
| 0 | 1.321.060 | 395.970 (30) |
| 18-29 | 207.060 | 30.330 (15) |
| 30-39 | 216.190 | 44.770 (21) |
| 40-49 | 325.770 | 91.370 (28) |
| 50-59 | 572.040 | 229.500 (40) |
| 1 | 19.340 | 5.310 (27) |
| 18-29 | 10.740 | 2.500 (23) |
| 30-39 | 4.390 | 1.170 (27) |
| 40-49 | 2.300 | 850 (37) |
| 50-59 | 1.910 | 800 (42) |
| **Voting proportions political movements (%)** | | |
| **Progressive liberal** | | |
| 0-0.1 | 138.940 | 35.900 (26) |
| 18-29 | 21.580 | 2.470 (11) |
| 30-39 | 21.920 | 3.620 (17) |
| 40-49 | 33.690 | 7.930 (24) |
| 50-59 | 61.750 | 21.880 (35) |
| 0.1-0.2 | 822.070 | 243.660 (30) |
| 18-29 | 121.630 | 17.430 (14) |
| 30-39 | 132.400 | 25.830 (20) |
| 40-49 | 202.900 | 55.330 (27) |
| 50-59 | 365.140 | 145.060 (40) |
| 0.2-0.3 | 239.540 | 77.610 (32) |
| 18-29 | 40.880 | 6.930 (17) |
| 30-39 | 41.340 | 10.100 (24) |
| 40-49 | 60.230 | 19.000 (32) |
| 50-59 | 97.090 | 41.580 (43) |
| 0.3-0.4 | 86.710 | 27.730 (32) |
| 18-29 | 22.980 | 4.120 (18) |
| 30-39 | 16.190 | 4.360 (27) |
| 40-49 | 18.740 | 6.320 (34) |
| 50-59 | 28.800 | 12.930 (45) |
| 0.4-0.5 | 10.580 | 3.600 (34) |
| 18-29 | 4.470 | 860 (19) |
| 30-39 | 1.580 | 530 (33) |
| 40-49 | 1.800 | 730 (41) |

| | | |
|---|---|---|
| 50-59 | 2.740 | 1.480 (54) |
| Unknown | 42.570 | 12.770 (30) |
| 18-29 | 6.250 | 1.010 (16) |
| 30-39 | 7.160 | 1.490 (21) |
| 40-49 | 10.730 | 2.910 (27) |
| 50-59 | 18.430 | 7.360 (40) |
| **Right-wing liberal** | | |
| 0-0.1 | 31.280 | 6.990 (22) |
| 18-29 | 6.380 | 950 (15) |
| 30-39 | 5.830 | 1.160 (20) |
| 40-49 | 7.170 | 1.510 (21) |
| 50-59 | 11.910 | 3.370 (28) |
| 0.1-0.2 | 507.570 | 138.040 (27) |
| 18-29 | 92.440 | 13.430 (15) |
| 30-39 | 85.360 | 16.820 (20) |
| 40-49 | 121.140 | 31.300 (26) |
| 50-59 | 208.630 | 76.480 (37) |
| 0.2-0.3 | 644.200 | 205.030 (32) |
| 18-29 | 95.730 | 14.590 (15) |
| 30-39 | 103.890 | 22.330 (21) |
| 40-49 | 159.890 | 47.420 (30) |
| 50-59 | 284.690 | 120.690 (42) |
| 0.3-0.4 | 114.430 | 38.310 (33) |
| 18-29 | 16.970 | 2.840 (17) |
| 30-39 | 18.310 | 4.120 (23) |
| 40-49 | 29.050 | 9.040 (31) |
| 50-59 | 50.100 | 22.300 (45) |
| 0.4-0.5 | 350 | 130 (37) |
| 18-29 | 30 | . |
| 30-39 | 30 | . |
| 40-49 | 100 | 30 (31) |
| 50-59 | 190 | 90 (47) |
| Unknown | 42.570 | 12.770 (30) |
| 18-29 | 6.250 | 1.010 (16) |
| 30-39 | 7.160 | 1.490 (21) |
| 40-49 | 10.730 | 2.910 (27) |
| 50-59 | 18.430 | 7.360 (40) |
| **Right-wing Christian** | | |
| 0-0.1 | 1.239.310 | 371.830 (30) |
| 18-29 | 202.040 | 30.580 (15) |
| 30-39 | 203.950 | 42.580 (21) |
| 40-49 | 303.260 | 85.590 (28) |
| 50-59 | 530.050 | 213.080 (40) |
| 0.1-0.2 | 33.720 | 10.140 (30) |

| | | |
|---|---|---|
| 18-29 | 5.400 | 760 (14) |
| 30-39 | 5.490 | 1.170 (21) |
| 40-49 | 8.210 | 2.270 (28) |
| 50-59 | 14.620 | 5.950 (41) |
| 0.2-0.3 | 14.650 | 4.000 (27) |
| 18-29 | 2.440 | 290 (12) |
| 30-39 | 2.420 | 420 (17) |
| 40-49 | 3.480 | 890 (26) |
| 50-59 | 6.310 | 2.410 (38) |
| 0.3-0.4 | 3.890 | 770 (20) |
| 18-29 | 640 | 50 (7) |
| 30-39 | 620 | 70 (11) |
| 40-49 | 950 | 170 (18) |
| 50-59 | 1.670 | 480 (29) |
| 0.4-0.5 | 2.890 | 820 (28) |
| 18-29 | 530 | 90 (17) |
| 30-39 | 430 | 100 (24) |
| 40-49 | 700 | 180 (26) |
| 50-59 | 1.230 | 450 (36) |
| 0.5-0.6 | 1.130 | 210 (19) |
| 18-29 | 170 | 10 (8) |
| 30-39 | 190 | 30 (16) |
| 40-49 | 250 | 50 (19) |
| 50-59 | 520 | 120 (23) |
| 0.7+ | 20 | . |
| 18-29 | . | . |
| 30-39 | . | . |
| 40-49 | . | . |
| 50-59 | . | . |
| 60-69 | . | . |
| 70-79 | . | . |
| 80+ | . | . |
| Unknown | 44.800 | 13.490 (30) |
| 18-29 | 6.570 | 1.060 (16) |
| 30-39 | 7.480 | 1.560 (21) |
| 40-49 | 11.210 | 3.060 (27) |
| 50-59 | 19.530 | 7.810 (40) |
| **Christian middle** | | |
| 0-0.1 | 509.040 | 144.260 (28) |
| 18-29 | 90.790 | 13.810 (15) |
| 30-39 | 87.100 | 18.140 (21) |
| 40-49 | 123.330 | 33.400 (27) |
| 50-59 | 207.820 | 78.910 (38) |
| 0.1-0.2 | 634.840 | 200.470 (32) |

| | | |
|---|---|---|
| 18-29 | 96.120 | 14.800 (15) |
| 30-39 | 101.380 | 21.550 (21) |
| 40-49 | 156.470 | 46.100 (29) |
| 50-59 | 280.870 | 118.030 (42) |
| 0.2-0.3 | 124.990 | 35.990 (29) |
| 18-29 | 19.820 | 2.690 (14) |
| 30-39 | 20.220 | 3.890 (19) |
| 40-49 | 30.540 | 8.050 (26) |
| 50-59 | 54.420 | 21.370 (39) |
| 0.3-0.4 | 25.590 | 7.030 (27) |
| 18-29 | 4.180 | 470 (11) |
| 30-39 | 4.200 | 790 (19) |
| 40-49 | 6.170 | 1.590 (26) |
| 50-59 | 11.050 | 4.190 (38) |
| 0.4-0.5 | 3.370 | 770 (23) |
| 18-29 | 630 | 60 (9) |
| 30-39 | 540 | 80 (16) |
| 40-49 | 840 | 170 (21) |
| 50-59 | 1.360 | 450 (33) |
| Unknown | 42.570 | 12.770 (30) |
| 18-29 | 6.250 | 1.010 (16) |
| 30-39 | 7.160 | 1.490 (21) |
| 40-49 | 10.730 | 2.910 (27) |
| 50-59 | 18.430 | 7.360 (40) |
| **Right-wing conservative** | | |
| 0-0.1 | 92.020 | 28.260 (31) |
| 18-29 | 23.880 | 4.210 (18) |
| 30-39 | 16.630 | 4.310 (26) |
| 40-49 | 20.240 | 6.450 (32) |
| 50-59 | 31.270 | 13.290 (42) |
| 0.1-0.2 | 539.020 | 170.960 (32) |
| 18-29 | 89.070 | 14.560 (16) |
| 30-39 | 90.160 | 20.770 (23) |
| 40-49 | 133.620 | 40.310 (30) |
| 50-59 | 226.160 | 95.320 (42) |
| 0.2-0.3 | 577.640 | 167.270 (29) |
| 18-29 | 86.380 | 11.730 (14) |
| 30-39 | 92.830 | 17.480 (19) |
| 40-49 | 141.910 | 37.820 (27) |
| 50-59 | 256.520 | 100.240 (39) |
| 0.3-0.4 | 83.030 | 20.810 (25) |
| 18-29 | 11.400 | 1.240 (11) |
| 30-39 | 12.940 | 1.790 (14) |
| 40-49 | 20.070 | 4.510 (22) |

| | | |
|---|---|---|
| 50-59 | 38.620 | 13.270 (34) |
| 0.4-0.5 | 3.470 | 770 (22) |
| 18-29 | 400 | 40 (10) |
| 30-39 | 520 | 60 (11) |
| 40-49 | 820 | 150 (19) |
| 50-59 | 1.740 | 520 (30) |
| 0.5-0.6 | 2.650 | 440 (16) |
| 18-29 | 420 | 30 (8) |
| 30-39 | 340 | 40 (10) |
| 40-49 | 690 | 70 (11) |
| 50-59 | 1.200 | 300 (25) |
| Unknown | 42.570 | 12.770 (30) |
| 18-29 | 6.250 | 1.010 (16) |
| 30-39 | 7.160 | 1.490 (21) |
| 40-49 | 10.730 | 2.910 (27) |
| 50-59 | 18.430 | 7.360 (40) |
| **Progressive left-wing** | | |
| 0-0.1 | 39.700 | 9.910 (25) |
| 18-29 | 6.880 | 750 (11) |
| 30-39 | 6.380 | 1.080 (17) |
| 40-49 | 9.560 | 2.140 (22) |
| 50-59 | 16.890 | 5.940 (35) |
| 0.1-0.2 | 488.360 | 154.080 (32) |
| 18-29 | 73.030 | 11.210 (15) |
| 30-39 | 77.810 | 16.370 (21) |
| 40-49 | 122.120 | 35.770 (29) |
| 50-59 | 215.400 | 90.730 (42) |
| 0.2-0.3 | 598.460 | 175.420 (29) |
| 18-29 | 92.520 | 13.330 (14) |
| 30-39 | 98.880 | 19.920 (20) |
| 40-49 | 147.360 | 40.430 (27) |
| 50-59 | 259.700 | 101.740 (39) |
| 0.3-0.4 | 150.660 | 43.560 (29) |
| 18-29 | 35.410 | 5.990 (17) |
| 30-39 | 26.510 | 6.240 (24) |
| 40-49 | 33.610 | 9.770 (29) |
| 50-59 | 55.140 | 21.560 (39) |
| 0.4-0.5 | 20.650 | 5.530 (27) |
| 18-29 | 3.710 | 540 (14) |
| 30-39 | 3.850 | 840 (22) |
| 40-49 | 4.710 | 1.190 (25) |
| 50-59 | 8.390 | 2.970 (35) |
| Unknown | 42.570 | 12.770 (30) |
| 18-29 | 6.250 | 1.010 (16) |

| | | |
|---|---|---|
| 30-39 | 7.160 | 1.490 (21) |
| 40-49 | 10.730 | 2.910 (27) |
| 50-59 | 18.430 | 7.360 (40) |
